# Self-supervised learning of accelerometer data provides new insights for sleep and its association with mortality

**DOI:** 10.1101/2023.07.07.23292251

**Authors:** Hang Yuan, Tatiana Plekhanova, Rosemary Walmsley, Amy C. Reynolds, Kathleen J. Maddison, Maja Bucan, Philip Gehrman, Alex Rowlands, David W. Ray, Derrick Bennett, Joanne McVeigh, Leon Straker, Peter Eastwood, Simon D. Kyle, Aiden Doherty

## Abstract

**Background:** Sleep is essential to life. Accurate measurement and classification of sleep/wake and sleep stages is important in clinical studies for sleep disorder diagnoses and in the interpretation of data from consumer devices for monitoring physical and mental well-being. Existing non-polysomnography sleep classification techniques mainly rely on heuristic methods developed in relatively small cohorts. Thus, we aimed to establish the accuracy of wrist-worn accelerometers for sleep stage classification and subsequently describe the association between sleep duration and efficiency (proportion of total time asleep when in bed) with mortality outcomes.

**Methods:** We developed and validated a self-supervised deep neural network for sleep stage classification using concurrent laboratory-based polysomnography and accelerometry data from three countries (Australia, the UK, and the USA). The model was validated within-cohort using subject-wise five-fold cross-validation for sleep-wake classification and in a three-class setting for sleep stage classification wake, rapid-eye-movement sleep (REM), non-rapid-eye-movement sleep (NREM) and by external validation. We assessed the face validity of our model for population inference by applying the model to the UK Biobank with 100,000 participants, each of whom wore a wristband for up to seven days. The derived sleep parameters were used in a Cox regression model to study the association of sleep duration and sleep efficiency with all-cause mortality.

**Findings:** After exclusion, 1,448 participant nights of data were used to train the sleep classifier. The difference between polysomnography and the model classifications on the external validation was 34.7 minutes (95% limits of agreement (LoA): −37.8 to 107.2 minutes) for total sleep duration, 2.6 minutes for REM duration (95% LoA: −68.4 to 73.4 minutes) and 32.1 minutes (95% LoA: −54.4 to 118.5 minutes) for NREM duration. The derived sleep architecture estimate in the UK Biobank sample showed good face validity. Among 66,214 UK Biobank participants, 1,642 mortality events were observed. Short sleepers (<6 hours) had a higher risk of mortality compared to participants with normal sleep duration (6 to 7.9 hours), regardless of whether they had low sleep efficiency (Hazard ratios (HRs): 1.69; 95% confidence intervals (CIs): 1.28 to 2.24) or high sleep efficiency (HRs: 1.42; 95% CIs: 1.14 to 1.77).

**Interpretation:** Deep-learning-based sleep classification using accelerometers has a fair to moderate agreement with polysomnography. Our findings suggest that having short overnight sleep confers mortality risk irrespective of sleep continuity.

**Funding:** This research has been conducted using the UK Biobank Resource under Application Number 59070. The UK Biobank received ethical approval from the National Health Service National Research Service (Ref 21/NW/0157). We would like to acknowledge the Raine Study participants and their families for their ongoing participation in the study and the Raine Study team for study coordination and data collection. We also thank the NHMRC for their long-term contribution to funding the study over the last 30 years. The core management of the Raine Study is funded by The University of Western Australia, Curtin University, Telethon Kids Institute, Women and Infants Research Foundation, Edith Cowan University, Murdoch University, The University of Notre Dame Australia and the Raine Medical Research Foundation. The 22-year Gen2 Raine Study follow-up was funded by NHMRC project grants 1027449 & 1044840. The data collection for the Pennsylvania dataset is funded, in part, by US National Institute of Health (NIMH) grant R21 MH103963 (MB).

HY, DB, and AD are supported by Novo Nordisk. RW and AD are supported by Health Data Research UK, an initiative funded by UK Research and Innovation, Department of Health and Social Care (England) and the devolved administrations, and leading medical research charities. AD is additionally supported by Swiss Re, Wellcome Trust [223100/Z/21/Z], and the British Heart Foundation Centre of Research Excellence (grant number RE/18/3/34214). DWR is supported by MRC programme grant MR/P023576/1; Wellcome Trust (107849/Z/15/Z). TP and AR are supported by the National Institute for Health Research (NIHR) Leicester Biomedical Research Centre and NIHR Applied Research Collaboration East Midlands (ARC EM). SDK is supported by the NIHR Oxford Health Biomedical Research Centre, Health Technology Assessment Programme, Efficacy and Mechanisms Evaluation Programme, Programme Grants for Applied Research, and the Wellcome Trust. The views expressed are those of the authors and not necessarily those of the NHS, the NIHR or the Department of Health.

Computational aspects of this research were funded from the National Institute for Health Research (NIHR) Oxford Biomedical Research Centre (BRC) with additional support from Health Data Research (HDR) UK and the Wellcome Trust Core Award [grant number 203141/Z/16/Z]. The views expressed are those of the authors and not necessarily those of the NHS, the NIHR or the Department of Health.

For the purpose of open access, the author has applied a CC-BY public copyright licence to any author accepted manuscript version arising from this submission.

**Research in context:** *Evidence before this study:* Sleep plays a crucial role in our mental and physical health. Nonetheless, much of our understanding of sleep relies on self-report sleep questionnaires, which are subject to recall bias. We searched on Web of Science, Medline, and Google Scholar from the database inception to June 23, 2023, using terms that included “wearable”, “actigraphy” or “accelerometer” in combination with “sleep stage” or “sleep classification”, and “polysomnography”. Existing studies have attempted to use machine learning to predict both sleep and sleep stages using accelerometry. However, prior methods were validated in populations of small sample sizes (n<100), making the prediction validity unclear. To date, no study has examined variations of accelerometer-derived sleep stage estimates in large population datasets with longitudinal disease outcomes.

*Added value of this study:* We showed that our deep-learning-based method improves sleep staging for wrist-worn accelerometers against the current state-of-the-art. We quantified the model uncertainty in a large multicentre dataset with 1,448 nights of concurrent raw accelerometry and polysomnography recordings. We further demonstrated that our sleep staging method could capture population differences concerning age, season, and other sociodemographic characteristics using a large health database. Shorter overnight sleep duration was associated with an increased risk of all-cause mortality after seven years of follow-up in groups with both low and high sleep efficiencies.

*Implications of all the available evidence:* This study helps clinicians to interpret sleep measurements from wearable sensors in routine care. Researchers can use derived sleep parameters in large-scale accelerometer datasets to advance our understanding of the association between sleep and population subgroups with different clinical characteristics. Our findings further suggest that having a short overnight sleep is a risky behaviour regardless of the sleep quality, which requires immediate public attention to fight the social stigma that having a short sleep is acceptable as long as one sleeps well.

## 1. Introduction

Sleep is essential to life and is structurally complex. Humans spend approximately one third of their lives asleep, yet sleep is hard to assess in free-living environments [1]. Our understanding of how sleep is associated with health and morbidity primarily draws on studies that use self-report sleep diaries, which capture the subjective experience [2]. However, sleep diaries have a low correlation with objective device-measured sleep parameters [3, 4]. The accepted standard for sleep measurement is laboratory-based polysomnography, which monitors sleep using a range of physical and physiological signals. However, polysomnography is not feasible for use at scale due to its high cost and technical complexity. Instead, wrist-worn accelerometers are more viable to deploy in large-scale epidemiological studies because of their portability and low user burden.

Despite the popularity of sleep monitoring in consumer and research-grade wristworn devices, sleep assessment algorithms are frequently proprietary and validated in small populations, making their measurement validity unclear [5, 6, 7, 8]. Methods for Sleep classification (i.e. defining periods of wake, NREM and REM sleep) primarily rely on hand-crafted spatiotemporal features such as device angle, which may not make full use of all the information in the signals. Hence, data-driven methods like deep learning could be advantageous. Furthermore, existing actigraphy-based sleep studies on large health datasets have only focused on the differentiation between sleep and wakefulness [9, 4, 10, 11] without evaluating variations in the stages of sleep.

We therefore set out to: (1) develop and internally validate an open-source novel deep learning method to infer sleep stages from wrist-worn accelerometers, (2) externally validate our proposed algorithm together with existing sleep staging bench-marks, and (3) investigate the association between device-measured overnight sleep duration and efficiency with all-cause mortality.

## 2. Methods

### 2.1. Study design and participants

In our multicentre cohort study, we developed and tested a sleep staging model for accelerometers (SleepNet) using a self-supervised deep recurrent neural network. We designed the model to classify each 30-second window of accelerometry data into one of the three sleep stages, wake, rapid-eye-movement sleep (REM), and non-rapid-eye movement sleep (NREM). Figure 1 illustrates the three main steps in our study: (1) feature extraction from unlabelled free-living data, (2) sleep staging model development, and (3) face validity assessment and health association analysis using the machine learning-estimated sleep parameters.

**Figure 1:**
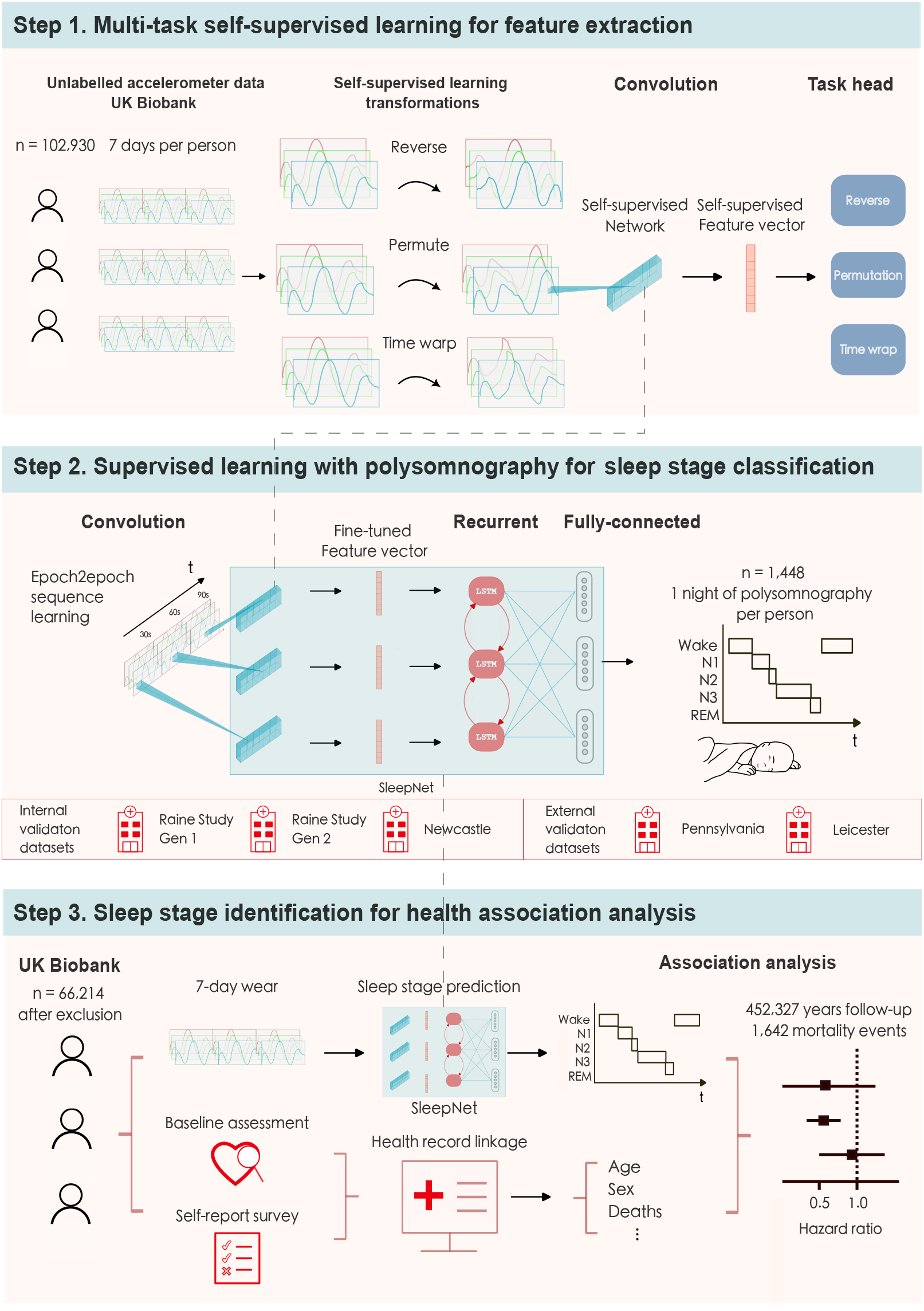
The SleepNet development pipeline: 1. We use multi-task self-supervised learning to obtain a feature extractor by learning from 700,000 person-days of tri-axial accelerometry data in the UK Biobank. 2. The pre-trained feature extractor was then fine-tuned with a deep recurrent network to train a sleep-stage classifier using polysomnography as the ground truth. 3. We deploy the sleep prediction model on the UK Biobank and investigate the association between device-measured sleep and mortality outcomes.

We used the UK Biobank accelerometry dataset [12] for two purposes: learning health-relevant accelerometer features to support the training of the sleep staging model and conducting the downstream health association analyses using the developed sleep staging model.

For sleep staging model development, internal validation consisted of two generations of participants from the Raine Study [13, 14] and a sleep patient population from the Newcastle cohort [15]. The Raine Study has followed up roughly 2900 children since 1989 in Australia. A subset of children (Raine Generation 2, Gen2) at the age of 22 and their parents (Raine Generation 1, Gen1) were invited to undergo one night of laboratory-based polysomnography at Western Australia’s Center for Sleep Science. The external validation consisted of two general populations from Leicester [16] and Pennsylvania [17]. Detailed population characteristics and inclusion criteria are listed in Supplementary Section 5.

### 2.2. Accelerometer devices and data preprocessing

Three different devices were used to collect the accelerometry for the included datasets, ActiGraph GT3X, Axivity AX3 and GENEActive Original accelerometers. The devices used have been shown to have a high inter-instrument agreement (> 80%) in derived sedentary and sleep-related time estimates in free-living environments [18]. As for device placement, we selected data from the dominant wrist where possible to be consistent with the UK Biobank protocol.

We used the Biobank Accelerometer Analysis Tool [19, 20] to preprocess all the data. The raw tri-axial accelerometry was first resampled into 30 Hz and clipped to ± 3 *g*. The accelerometry sequence was then divided into consecutive 30-second windows. We considered stationary periods (x/y/z sd < 13 m*g*) with a duration greater than 60 minutes as non-wear [12]. We further excluded the data that could not be parsed, had unrealistic high values (> 200 m*g*), or were poorly calibrated.

### 2.3. Ascertainment of sleep stages via polysomnography

The gold-standard, laboratory-based polysomnography sleep label was aligned with its concurrent accelerometer data as the model ground truth. The polysomnography labels were scored according to the American Academy of Sleep Medicine (AASM) protocol [21], which divided sleep into five categories: wake, REM, and NREM I, II, and III. In total, 1,157,913 (~10,000 hours) sleep windows were used to train the network. The sleep stage distributions were similar across all the datasets except for the Newcastle cohort, which had a greater proportion of wakefulness than the others (Supplementary Figure 1).

### 2.4. Deep learning analysis of sleep stages from wrist-worn accelerometers

A deep recurrent neural network (SleepNet) was trained to classify the sleep stages for every 30-second window of tri-axial accelerometry data. The SleepNet has three components: a ResNet-17 V2 [22] with 1D convolution for feature extraction, a bi-directional Long-Short-Term-Memory (LSTM) network for temporal dependencies learning [23], and two fully-connected (FC) layers for sleep stage prediction. During training, we provided the SleepNet with five-stage polysomnography labels (wake, REM, and NREM I, II, III). When evaluating the model, we collapsed all the NREM stages into one class for classification (wake/REM/NREM). Similarly, we collapsed all the REM and NREM stages together to classify wake vs sleep.

The SleepNet was pre-trained using multi-task self-supervision on the UK Biobank to learn features of human motion dynamics [24]. Multi-task self-supervision automatically extracts the features relevant to motion by learning to discriminate different spatiotemporal transformations applied to the unlabelled 700,000 person-days of data. Self-supervised pre-training has been shown to help classify human activity recognition not just in healthy but clinical populations [25]. See Supplementary Section 6 for further details of the model development.

For internal validation, we used subject-wise five-fold cross-validation on the Raine Gen2, Raine Gen1, and Newcastle cohorts. For external validation, we trained the SleepNet on all the internal datasets and then evaluated its performance on the Leicester and Pennsylvania cohorts. We compared the SleepNet performance with a random forest model that used the hand-crafted spatiotemporal features [20, 26]. The random forest feature definitions are listed in Supplementary Table 2.

We reported the staging performance in both subject-wise and epoch-to-epoch fashion. Three-class and five-class confusion matrices were plotted for both internal and external validation. Since Cohen Kappa, F1 scores, and balanced accuracies (Supplementary Table 3) are less influenced by class imbalance, they were used to evaluate the overall model. To assess the relationship between the model performance and population characteristics, we stratified the subject-wise sleep staging performance by age, sex, employment status, income level, body mass index (BMI), presence and severity of sleep apnea using the apnea-hypopnea index (AHI), existing sleep disorders, and neurological disorders where available.

Finally, we evaluated the agreement between summary sleep parameters per each night derived from our deep learning method and polysomnography via Bland-Altman plots for the following sleep parameters: total sleep duration, sleep efficiency (proportion of total time asleep when in bed), time awake after sleep onset (WASO), REM duration, NREM duration, REM ratio, NREM ratio. Supplementary Table 4 entails the sleep parameter definitions and their calculations.

### 2.5. Measurements of sleep in 100,000 UK Biobank participants

We obtained the sleep architecture estimates on the UK Biobank by applying SleepNet on the longest overnight sleep windows. Since no concurrent sleep diaries were collected in the UK Biobank, we used a random forest model trained on sleep diaries with Hidden Markov Models smoothing to first obtain time in bed [19, 20]. The random forest model achieved 90%+ precision and recall for detecting sleep windows in 152 free-living participants with sleep diaries that asked two questions: “what time did you first fall asleep last night?” and “what time did you wake up (eyes open, ready to get up)?” [20]. We used the sleep window output from the random forest model as a proxy for the time in bed. We then merged any time in bed windows within 60 minutes of one another [27]. Finally, we applied the SleepNet on the longest window over each noon-to-noon interval to estimate the overnight sleep duration. The difference between overnight and total sleep duration is that total sleep duration is a sleep parameter used to assess the agreement between our SleepNet output and polysomnography for model validation. Overnight sleep duration refers to the estimate for the amount of sleep one obtains for a noon-to-noon interval in a free-living environment using a random forest model for sleep window detection and the SleepNet for sleep stage identification.

We simulated the effects of random missing data on the participants that had no missing data across seven-days to determine the minimum wear time required for stable weekly sleep parameter estimates (Supplementary Section 7.2). We found that a minimum of 22 hours of wear time per day for at least three days were required to ensure the intra-class correlation was greater than 0.75 between the weekly average sleep duration from incomplete and perfect wear data. Moreover, we tried to mitigate the weekend effect by only including the participants who had at least one weekday and one weekend day during the device wear. Shift workers and participants whose data had daylight saving cross-overs were also excluded, as circadian disruption is not the focus of our paper.

Descriptive analyses were performed on the device-measured sleep parameters in the UK Biobank to quantify variations by age, sex, device-measured physical activity level, self-reported chronotype and insomnia symptoms. Estimated marginal means, adjusted for age and sex, were also calculated for different self-rated health groups and self-reported insomnia symptoms.

### 2.6. Health association analysis

The associations of overnight sleep duration and sleep efficiency with incident mortality were assessed using Cox proportional hazards regression. All-cause mortality was determined using death registry data (obtained by UK Biobank from NHS Digital for participants in England and Wales and from the NHS Central Register, National Records of Scotland, for participants in Scotland). Participants were censored at the earliest of UK Biobank’s record censoring date for mortality data (2021-09-30 for participants in England and Wales and 2021-10-31 for participants in Scotland, with country assigned based on baseline assessment centre). Cox models used age as the timescale, and the main analysis was adjusted for sex, ethnicity, Townsend Deprivation Index, educational qualifications, smoking status, alcohol consumption, and overall activity. See Supplementary Section 7.1 for the full specification of the analysis.

## 3. Results

### 3.1. Comparison to polysomnography

After preprocessing, 1,395 participants were included in the internal validation, and 53 participants were included in the external validation. Our proposed deep recurrent neural network (SleepNet) pre-trained with self-supervision achieved the best performance when compared with other baseline models that used hand-crafted features (Supplementary Table 6).

On the internal validation, SleepNet had a mean bias of 8.9 minutes (95% limits of agreement (LoA): −89.0 to 106.9 minutes) for total sleep duration, −18.7 minutes (95% LoA: −130.9 to 93.6 minutes) for REM duration, and 27.6 minutes (95% LoA: −100.6 to 155.8 minutes) for NREM duration (Figure 2). In comparison, on the external validation, the mean bias was 34.7 minutes (95% LoA: −37.8 to 107.2 minutes) for total sleep duration, −2.6 minutes (95% LoA: −68.4 to 73.6 minutes) for REM duration, and 32.1 minutes (95% LoA: −54.4 to 118.5 minutes) for NREM duration. Overall, our model tends to underestimate REM and short sleep and overestimate NREM and long sleep. Supplementary Figures 5 to 10 depict the agreement assessments for other sleep parameters on the individual cohorts.

**Figure 2:**
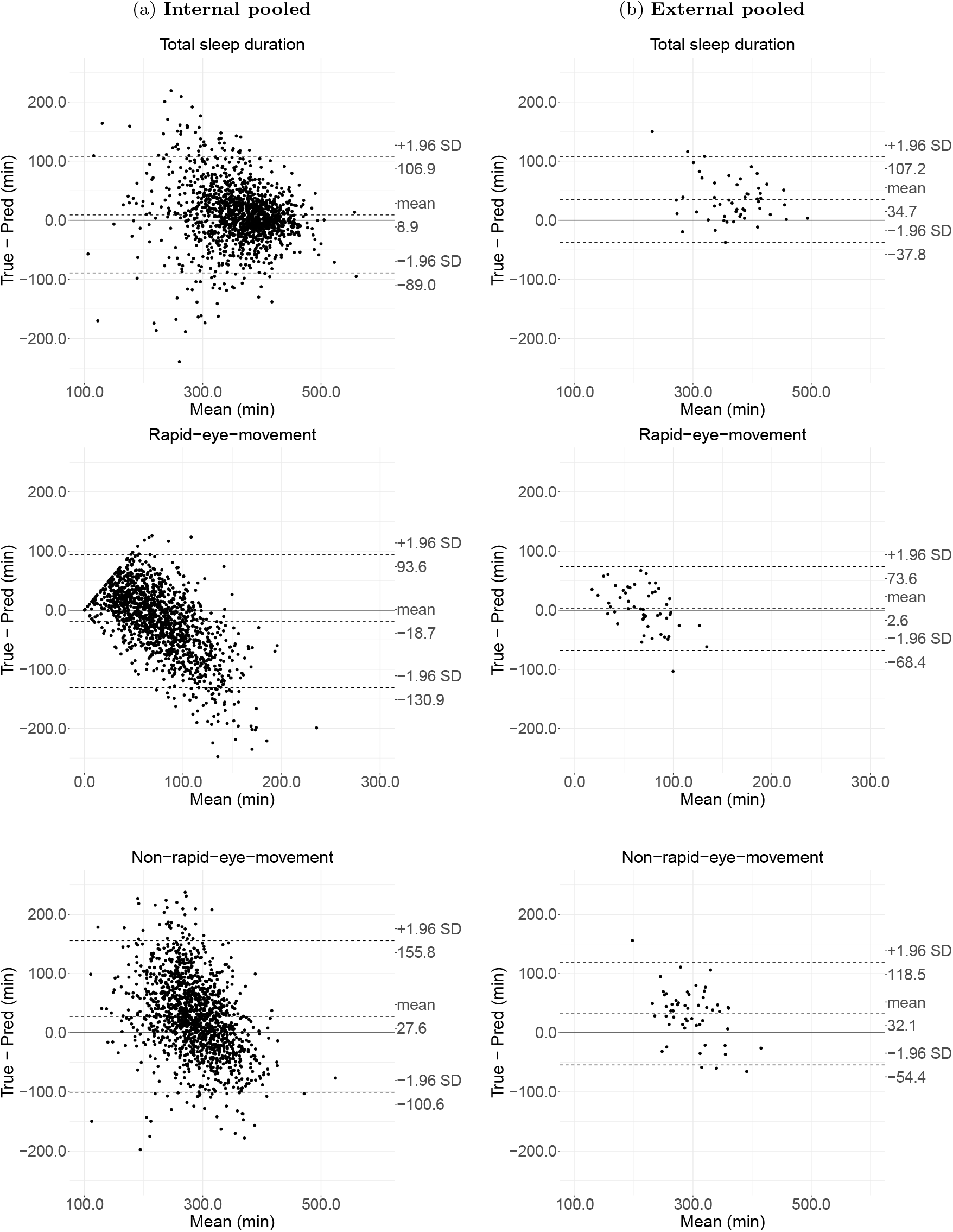
Agreement assessment via Bland-Atman plot for total sleep duration, rapid eye movement sleep (REM) duration, and non-rapid eye movement sleep (NREM) duration on internal and external validation. The internal validation consists of 1,373 polysomnography nights from the Raine Study and the Newcastle cohort, whereas the external validation consists of 53 polysomnography nights from the Leicester and Pennsylvania cohorts.

The subject-wise performance for both the internal and external validation using the pre-trained SleepNet is shown in Supplementary Table 7. On the pooled internal validation, our model obtained an F1 of 0.75 ± 0.1 in the two-class setting (sleep/wake) and an F1 of 0.57 ± 0.11 in the three-class setting (wake/REM/NREM). The agreement decreased slightly on the external validation with an F1 of 0.67 ± 0.11 in the two-class setting (sleep/wake) and an F1 of 0.52 ± 0.10 in the three-class setting (wake/REM/NREM). In the Newcastle cohort, for the sleep/wake classification, sensitivity decreased and specificity increased in participants with sleep disorders. No obvious difference was observed in both Raine Gen1 and Gen2 cohorts when the participants were stratified by sex, BMI, AHI, and sleep disorder conditions.(Supplementary Table 8–10).

To classify any given window in an epoch-by-epoch fashion, the SleepNet achieved a Kappa score of 0.39 on the internal validation set and a Kappa score of 0.32 on the external validation set in the three-class setting (Supplementary Figure 11). Cohort-specific confusion matrices can be found in Supplementary Figures 12–15. Supplementary Figure 16 visualizes a one-night sample actigram, its ground-truth polysomnography labels, and SleepNet predictions. We used SleepNet to generate all the sleep parameters for the rest of the paper.

### 3.2. Face validity in the UK Biobank

Before deploying the SleepNet on the UK Biobank, we excluded participants with unusable accelerometer data and participants with missing covariates in the descriptive analysis. We further excluded participants with any prior hospitalisation for cardiovascular disease or cancer in the association analysis (Supplementary Figure 17). In sum, 66,214 participants were included in the final analysis.

Table 1 describes the variations in overnight sleep duration, REM and NREM durations, and sleep efficiency across population subgroups in the UK Biobank. Older participants generally slept longer with higher sleep efficiency. Females had a longer overnight sleep duration and NREM but a shorter REM than males. Participants with better self-rated health had longer sleep duration and higher sleep efficiency than those with poor self-rated health. Sleep efficiency was relatively stable across different seasons and days of the week. The correlation coefficients between device-measured sleep parameters during accelerometer wear and self-reported total sleep duration at baseline assessment were all below 0.25 (Supplementary Figure 18). The distributions of device-measured overnight sleep duration tend to have a greater variability for participants who self-reported to have less than 5 or greater than 10 hours of total sleep duration (Supplementary Figure 19). Overall, sleep stage distribution was similar for males and females aged between 45 and 75, with NREM sleep fluctuating around 5 hours and REM sleep fluctuating around 2.5 hours per night (Supplementary Figure 20). No major differences were seen between females and males.

**Table 1:** Overall sleep parameters by participant characteristics in the UK Biobank (mean ± SD) for overnight sleep duration, non-rapid-eye-movement sleep (NREM), rapid-eye-movement sleep (REM), and sleep efficiency.

| Characteristics | n (%) | Overnight sleep<br>h/day | NREM<br>h/day | REM<br>h/day | Sleep efficiency<br>% |
| --- | --- | --- | --- | --- | --- |
| Overall | 66214 (100.0) | 7.5±1.0 | 5.0±1.0 | 2.5±0.9 | 87.9±4.9 |
| Age, year |  |  |  |  |  |
| 40-49 | 6115 (9.2) | 7.4±0.9 | 4.7±0.9 | 2.6±0.9 | 87.9±4.7 |
| 50-59 | 20130 (30.4) | 7.4±0.9 | 4.9±1.0 | 2.5±0.9 | 87.7±4.9 |
| 60-69 | 29198 (44.1) | 7.5±1.0 | 5.0±1.0 | 2.5±0.9 | 88.0±4.9 |
| 70-79 | 10771 (16.3) | 7.5±1.0 | 5.0±1.0 | 2.5±0.9 | 88.2±5.0 |
| Sex |  |  |  |  |  |
| Female | 38525 (58.2) | 7.6±0.9 | 5.2±1.0 | 2.4±0.9 | 88.3±4.7 |
| Male | 27689 (41.8) | 7.3±1.0 | 4.7±0.9 | 2.7±0.9 | 87.4±5.1 |
| Ethnicity |  |  |  |  |  |
| Non-white | 2003 (3.0) | 7.0±1.1 | 4.8±1.0 | 2.2±0.9 | 86.3±5.6 |
| White | 64211 (97.0) | 7.5±0.9 | 5.0±1.0 | 2.5±0.9 | 88.0±4.9 |
| Physical activity level |  |  |  |  |  |
| low < 24.08 mg | 22058 (33.3) | 7.7±1.1 | 5.1±1.0 | 2.5±1.0 | 87.2±5.4 |
| Medium 24.08-30.42 mg | 22072 (33.3) | 7.5±0.9 | 5.0±1.0 | 2.5±0.9 | 88.1±4.7 |
| High > 30.42 mg | 22084 (33.4) | 7.3±0.9 | 4.8±0.9 | 2.5±0.9 | 88.5±4.5 |
| Smoking status |  |  |  |  |  |
| Never smoker | 38930 (58.8) | 7.5±0.9 | 5.0±1.0 | 2.5±0.9 | 88.0±4.8 |
| Ex-smoker | 22870 (34.5) | 7.5±1.0 | 5.0±1.0 | 2.5±0.9 | 88.0±4.9 |
| Current smoker | 4414 (6.7) | 7.3±1.0 | 5.0±1.0 | 2.3±0.9 | 87.4±5.5 |
| Alcohol consumption |  |  |  |  |  |
| Never drinker | 3607 (5.4) | 7.4±1.1 | 5.0±1.0 | 2.4±0.9 | 87.4±5.4 |
| < 3 times per week | 30074 (45.4) | 7.5±1.0 | 5.0±1.0 | 2.5±0.9 | 87.7±5.0 |
| 3+ times per week | 32533 (49.1) | 7.5±0.9 | 4.9±1.0 | 2.5±0.9 | 88.2±4.7 |
| Education |  |  |  |  |  |
| School leaver | 14648 (22.1) | 7.6±1.0 | 5.1±1.0 | 2.5±0.9 | 87.5±5.1 |
| Further education | 21700 (32.8) | 7.5±1.0 | 5.0±1.0 | 2.5±0.9 | 87.8±5.0 |
| Higher education | 29866 (45.1) | 7.4±0.9 | 4.9±1.0 | 2.5±0.9 | 88.2±4.7 |
| Townsend Deprivation Index |  |  |  |  |  |
| Least deprived (<-3.8) | 16552 (25.0) | 7.5±0.9 | 5.0±1.0 | 2.6±0.9 | 88.1±4.8 |
| Second least deprived<br>(-3.8 to -2.5) | 16554 (25.0) | 7.5±0.9 | 5.0±1.0 | 2.6±0.9 | 88.0±4.8 |
| Second most deprived<br>(-2.5 to -0.2) | 16552 (25.0) | 7.5±1.0 | 5.0±1.0 | 2.5±0.9 | 87.9±4.9 |
| Most deprived (> -0.2) | 16556 (25.0) | 7.4±1.0 | 5.0±1.0 | 2.4±0.9 | 87.8±5.1 |
| BMI |  |  |  |  |  |
| <18.5, underweight | 397 (0.6) | 7.5±1.0 | 5.1±1.0 | 2.5±0.9 | 89.1±4.7 |
| 18.5-24.9, normal | 26759 (40.4) | 7.6±0.9 | 5.0±1.0 | 2.6±0.9 | 88.4±4.6 |
| 25-29.9, overweight | 26920 (40.7) | 7.5±1.0 | 4.9±1.0 | 2.5±0.9 | 87.8±4.9 |
| 30+, obese | 12138 (18.3) | 7.3±1.1 | 5.0±1.0 | 2.3±0.9 | 87.1±5.4 |
| Employment |  |  |  |  |  |
| Employed | 41640 (62.9) | 7.4±0.9 | 4.9±1.0 | 2.5±0.9 | 87.9±4.8 |
| Not employed | 24574 (37.1) | 7.6±1.0 | 5.1±1.0 | 2.5±0.9 | 88.0±5.0 |
| Self-rated health |  |  |  |  |  |
| Poor | 1282 (1.9) | 7.4±1.3 | 5.0±1.1 | 2.3±1.0 | 87.0±6.0 |
| Fair | 9162 (13.8) | 7.4±1.1 | 5.0±1.0 | 2.4±0.9 | 87.3±5.3 |
| Good | 40120 (60.6) | 7.5±0.9 | 5.0±1.0 | 2.5±0.9 | 87.9±4.9 |
| Excellent | 15650 (23.6) | 7.5±0.9 | 4.9±1.0 | 2.6±0.9 | 88.4±4.6 |
| Day |  |  |  |  |  |
| Weekday | 66214 (100.0) | 7.4±1.0 | 4.9±1.0 | 2.5±0.9 | 88.0±5.2 |
| Weekend | 66214 (100.0) | 7.7±1.2 | 5.1±1.2 | 2.6±1.1 | 87.8±6.2 |
| Wear season |  |  |  |  |  |
| Spring | 14717 (22.2) | 7.5±0.9 | 4.9±1.0 | 2.5±0.9 | 87.9±4.9 |
| Summer | 18203 (27.5) | 7.4±0.9 | 4.9±1.0 | 2.4±0.9 | 88.2±4.8 |
| Autumn | 18682 (28.2) | 7.5±1.0 | 5.0±1.0 | 2.5±0.9 | 87.9±4.9 |
| Winter | 14612 (22.1) | 7.6±1.0 | 5.0±1.0 | 2.6±0.9 | 87.7±5.0 |

We found expected sleep-wake patterns in population subgroups. For example, timing of the sleep opportunity for participants with a self-reported “morning” chronotype was about one hour earlier when compared with those that had a self-reported “evening“ chronotype (Figure 3a). We saw similar but shorter phase advance (~30 mins) in participants who were most physically active compared to the participants that were least physically active (Figure 3b). When comparing groups that had a history of self-reported insomnia symptoms versus those who did not, we found that participants with a history of insomnia symptoms were less likely to be in REM sleep on average during the overnight sleep window (Figure 3d and Figure 3c). Participants with a history of self-reported insomnia symptoms tended to have a longer overnight sleep duration but with a lower sleep efficiency (Supplementary Figure 21). The sleep architecture for different population subgroups were similar between weekdays and weekends, with a slight phase delay over the weekend (Supplementary Figure 22).

**Figure 3:**
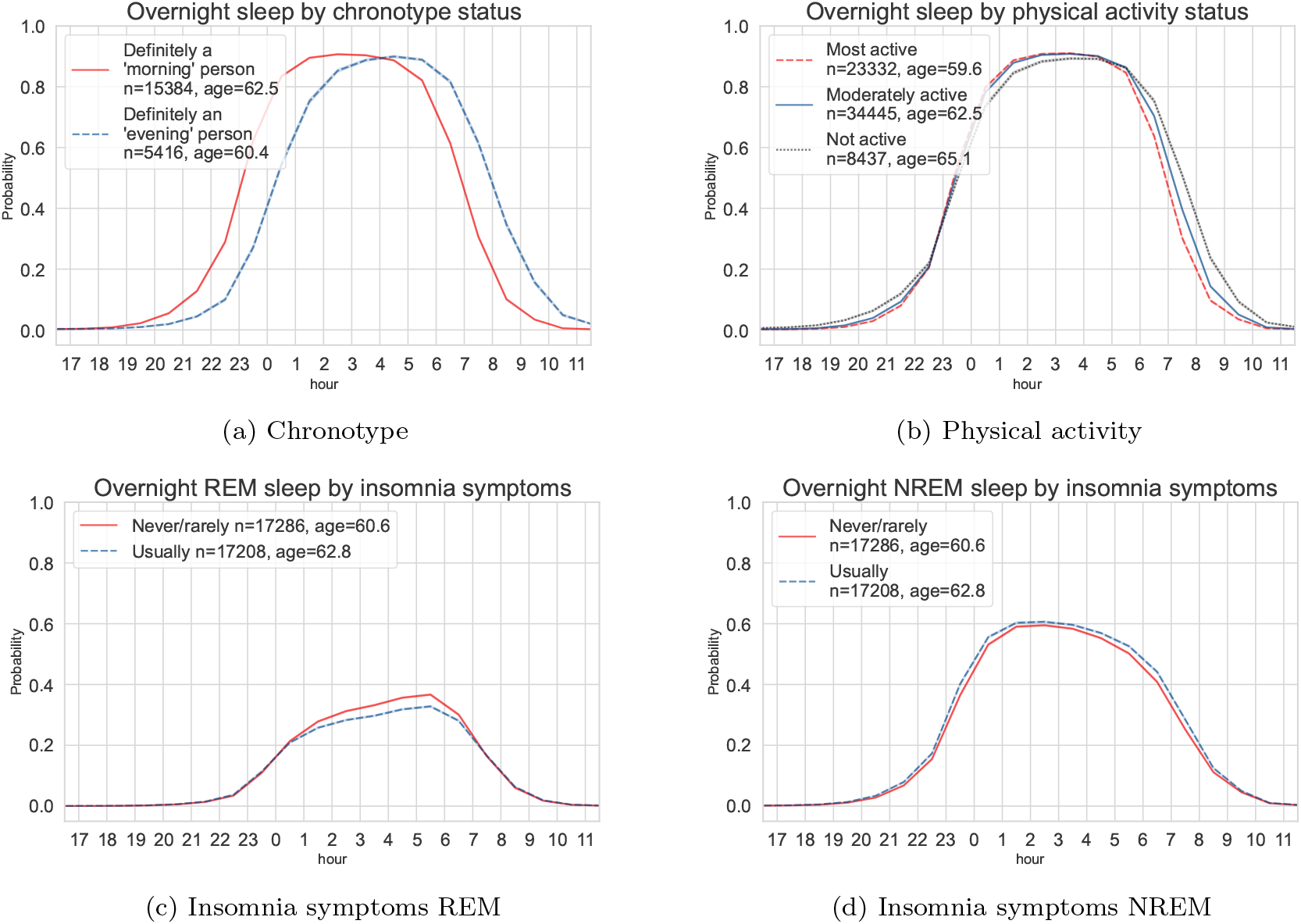
Device-measured sleep probability trajectories throughout the day for the UK Biobank participants. Top: variations of the average overnight sleep probability for the participants with self-reported “morning” and “evening” chronotype (a) and the overnight sleep distributions across thirds of device-measured physical activity level (b). Bottom: variations of the average REM (c) and NREM (d) probability in participants with a history of self-reported insomnia symptoms versus those without. REM: rapid-eye-movement sleep; NREM: non-rapid-eye-movement sleep.

### 3.3. Association with all-cause mortality

Over 452,327 years of the follow-up, 1,642 mortality events among 66,214 participants were observed. Short sleepers (<6 hours) had a higher risk of mortality in groups of low sleep efficiency (Hazard ratios (HRs): 1.69; 95% confidence intervals (CIs): 1.28 to 2.24) and high sleep efficiency (HRs: 1.42; 95% CIs: 1.14 to 1.77) compared to participants with normal sleep duration (6 to 7.9 hours, Figure 4). The risk of all-cause mortality appeared to decrease linearly as sleep efficiency increased. However, a non-linear association was observed in the association for overnight sleep duration (Supplementary Figure 23). When further adjusted for BMI, associations of overnight sleep duration and sleep efficiency with all-cause mortality were slightly attenuated (Supplementary Figure 24–25). Longer overnight sleep duration was not founded to have a higher risk than the reference group in both the main (Supplementary Figure 23) and sensitivity analysis (Supplementary Figure 26).

**Figure 4:**
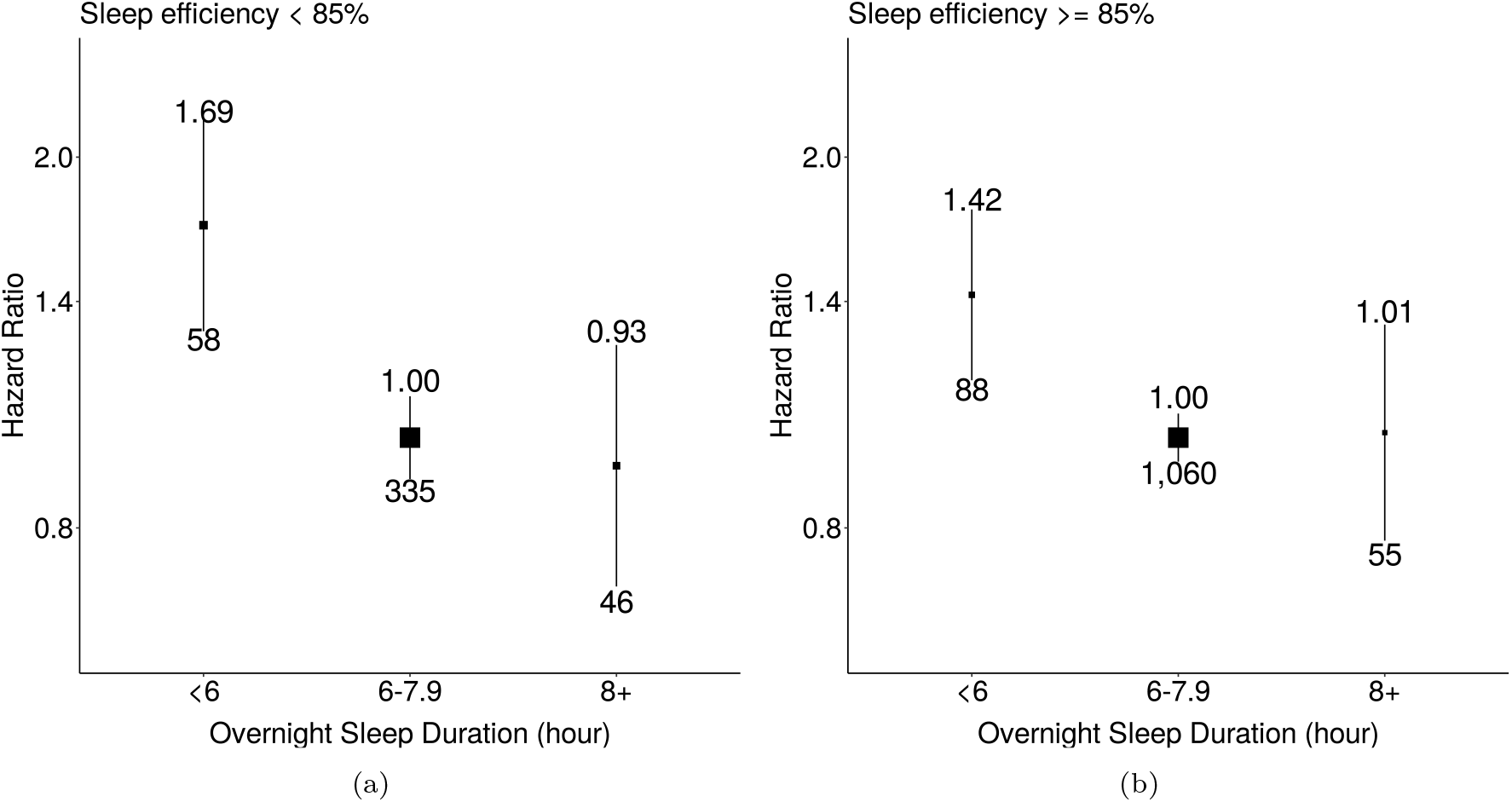
Associations of overnight sleep duration with all-cause mortality for groups with low and high sleep efficiency. The model used 1,642 events among 62,214 participants. We used age as the timescale and adjusted for sex, ethnicity, Townsend Deprivation Index of baseline address (split by quarter in the study population), educational qualifications, smoking status, alcohol consumption (Never, <3 times/week, 3+ times/week), overall activity (measured in milli-gravity units). Areas of squares represent the inverse of the variance of the log risk. The I bars denote the 95% confidence interval for the floated risks.

## 4. Discussion

We have developed, and internally and externally validated a deep-learning method to characterise sleep architecture from a wrist-worn accelerometer with competitive performance against 1,448 nights of laboratory-based polysomnography recordings. When applying our developed method in the UK Biobank in an epidemiological analysis of 66,214 participants, we found that shorter sleep time was associated with an increased risk of all-cause mortality individually regardless of sleep continuity, indexed by sleep efficiency. Our open-source algorithm and the inferred sleep parameters will open the door to future studies on sleep and sleep architecture using large-scale accelerometer databases.

Our novel self-supervised deep learning sleep staging method outperformed existing baseline methods that rely on hand-crafted features. The inferred sleep architecture estimates had a fair agreement (κ = 0.39) with the polysomnography ground truth on the internal validation [28]. Unlike previous work in sleep classification methods that depended on hand-crafted features [26, 29], our proposed method automatically extracted the features using self-supervision, hence removing the need for manual engineering. Even for sleep/wake classification, SleepNet achieved comparable results to a systematic evaluation of eight state-of-the-art sleep algorithms [8] in the Newcastle dataset. However, our work offers a more robust evaluation and identifies the upper limit of using accelerometry for sleep classification by developing a model with one of the largest multicentre datasets with polysomnography ground truth, at least ten times the size of existing studies.

In the subsequent epidemiological analysis, we found a clear association between short overnight sleep duration with increased risk of all-cause mortality in both good and poor sleepers defined by sleep efficiency. Short overnight sleep duration has been linked with mortality outcomes in self-report and actigraphy-based studies [30, 31]. However, few studies have investigated the joint effect of sleep duration and efficiency. One recent study has suggested that participants with short and long total sleep time had an increased risk after accounting for sleep efficiency [32]. However, our analysis did not find that long overnight sleep duration was associated with increased risk, potentially because we did not include daytime naps in our measurement of overnight sleep duration. Daytime napping has been found to be associated with an increased risk of cardiovascular events and deaths in those with longer nighttime sleep [33]. We did not find a U-shape association between device-measured sleep and mortality that has been suggested by other smaller studies [30]. Instead, our data are supportive of adverse associations with short sleep duration only, which is concordant with pre-clinical human and animal studies [34].

This study has several strengths, including the analysis of sleep architecture in a large, prospective Biobank with longitudinal follow-up. Compared with self-reported sleep questionnaires that only captured sleep duration to the nearest hour, actigraphy-based methods like ours can provide more fine-grained sleep duration and efficiency estimates. The extensive multicentre evaluation of the sleep classification allowed for the characterisation of the measurement uncertainty and a less biased interpretation of the health association analysis. Sleep stage identification from actigraphy is highly challenging, especially for wake periods in bed that are not characterised by wrist movement. With the proposed SleepNet, we could obtain sleep architecture estimates for population health inference after evaluating the face validity of the sleep parameters in the UK Biobank. While future work might improve sleep staging performance by incorporating additional physiological signals, such as electrocardiogram, to improve sleep staging performance, multi-modal sensor signals are not yet available for population-scale studies with longitudinal follow-up beyond a few years [35]. Despite our best efforts to include diverse validation cohorts from different centres, the included datasets mainly consist of healthy populations from a Caucasian ethnic background. Validation in populations with chronic diseases and different ethnic backgrounds would aid in quantifying the measurement uncertainty. In this work, we have developed and validated an open-source sleep staging method that substantially improves the ability to measure sleep characteristics with wrist-worn accelerometers in large biomedical datasets. Using the sleep parameters generated by our model, we demonstrated that shorter overnight sleep was associated with a higher risk of all-cause mortality in both good and poor sleepers. Our proposed method provides the community with a rich set of new measurements to study how sleep parameters are longitudinally associated with clinical outcomes.

## Data sharing

The data for the Newcastle cohort is available from direct download via https://zenodo.org/record/1160410#.Y-O65i-l1qs. The data for other cohorts can be requested by contacting the corresponding host institute. All the sleep staging models and analysis scripts are freely available for academic use on GitHub: https://github.com/OxWearables/asleep.

## Contributions

HY, KM, JM, LS, PE, SD, and AD conceptualised and designed the study. TP, MB, PG, AR, JM, LS, and PS did the data curation of the accelerometers and polysomnography data. HY, TP, and RW did the formal analysis and validation. DB, SK and AD provided supervision to HY and RW. HY wrote the manuscript, and all the authors contributed to the review & editing process. HY and RW had direct access to the summary statistics and verified the findings.

## Data Availability

The data for the Newcastle cohort is available from direct download via https://zenodo.org/record/1160410#.Y-O65i-l1qs. The data for other cohorts can be requested by contacting the corresponding host institute.

https://github.com/OxWearables/asleep

## Acknowledgments

We would like to thank Andrew Creagh, Angel Wong, Scott Small, and Alaina Shreves for their input on the revision of this manuscript. We would also like to thank Andrew Creagh for his feedback in creating the graphic illustrations.

## Supplements

**Table 1:**
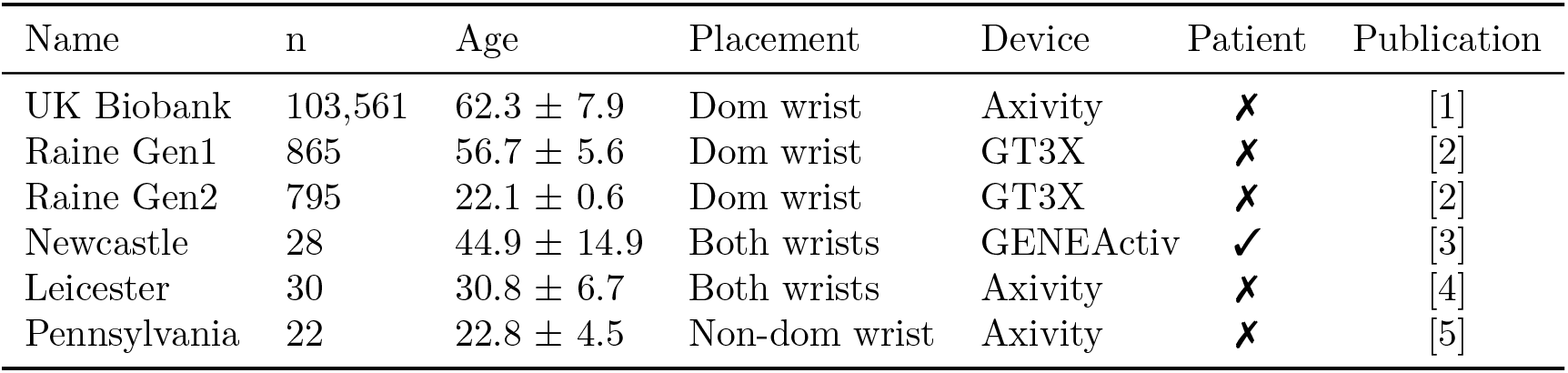
Characteristics of the datasets used for internal validation, external validation and health association analyses. “Patient” indicates whether a cohort consists of sleep patients in a clinic.

| Name | n | Age | Placement | Device | Patient | Publication |
| --- | --- | --- | --- | --- | --- | --- |
| UK Biobank | 103,561 | $62.3 \pm 7.9$ | Dom wrist | Axivity | ✗ | [1] |
| Raine Gen1 | 865 | $56.7 \pm 5.6$ | Dom wrist | GT3X | ✗ | [2] |
| Raine Gen2 | 795 | $22.1 \pm 0.6$ | Dom wrist | GT3X | ✗ | [2] |
| Newcastle | 28 | $44.9 \pm 14.9$ | Both wrists | GENEActiv | ✓ | [3] |
| Leicester | 30 | $30.8 \pm 6.7$ | Both wrists | Axivity | ✗ | [4] |
| Pennsylvania | 22 | $22.8 \pm 4.5$ | Non-dom wrist | Axivity | ✗ | [5] |

### 5. Datasets

#### Raine Study

The Raine Study has followed up roughly 2900 children since 1989 in Australia. A subset of children (Raine Gen2, 50% females) at the age of 22 and their parents (Raine Gen1, 57% females) were invited to undergo one night of laboratory-based polysomnography at Western Australia’s Center for Sleep Science [2, 6]. Every participant was instructed to wear an ActiGraph GT3X device on the dominant wrist. Earlier GT3X firmware would enter an idle mode to save the battery when no sufficient movement was detected, so we only included participants with no missing data for the Raine Gen2 cohort.

#### Newcastle

The Newcastle dataset recruited 28 adult patients (39% females) for a one night laboratory-based polysomnography assessment in Newcastle upon Tyne, UK, as part of their routine clinical visit [3]. During the polysomnography recording, the participants wore two GENEActive devices, one on each wrist. The sampling frequency for the wristbands was set to 85.7 Hz.

#### Leicester

Thirty healthy volunteers (63% females and 73% white) wore three devices: GENEActive, Axivity AX3, and ActiGraph GT9X on each wrist during one night of laboratory-based polysomnography assessment [4]. The relative position of the devices was randomly allocated for each participant. The devices were set to record at 100 Hz. During the lab visit, when the participants wished to go to bed, the recording was started. The sleep episodes usually ended between 6 am and 7 am the following morning. We cleaned up the recording sessions such that every recording would start from ”light off” and end at ”light off” to ensure comparability.

**Figure 1:**
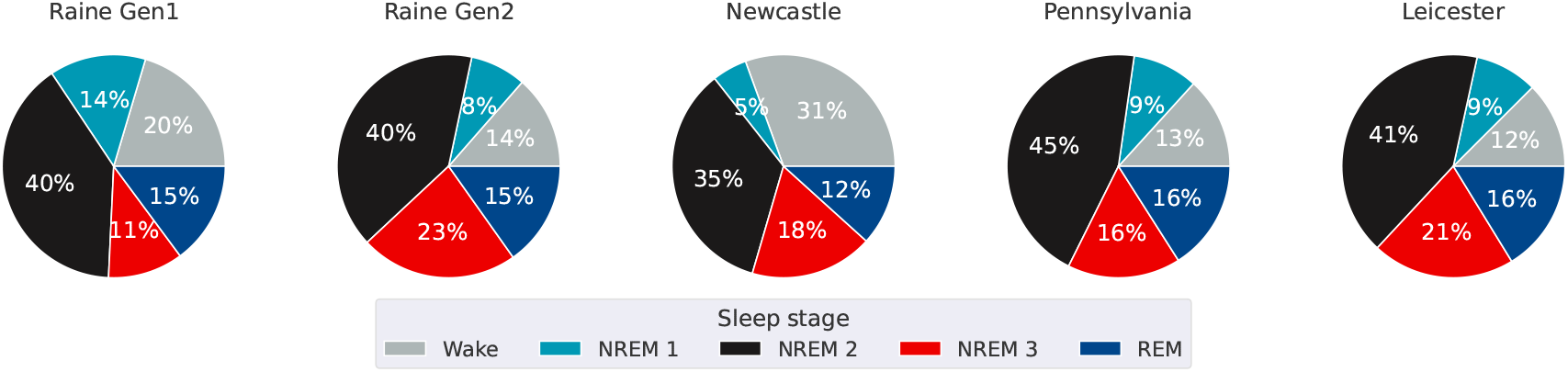
Sleep stage distribution for all the datasets used.

#### Pennsylvania

The Pennsylvania dataset consists of 22 healthy sleepers who had one-night of laboratory-based polysomnography assessment at the University of Pennsylvania Center for sleep [5]. The participants were asked to wear an Axivity device on the non-dominant wrist during the polysomnography session.

#### UK Biobank

The UK Biobank is a longitudinal cohort study that recruited 500,000 adults from the UK [7]. A subset of the participants was invited to wear an Axivity device on the dominant wrist for one week in a free-living environment [1]. The sampling rate was set to 100 Hz. Roughly 100,000 participants (56% females) consented and participated in the accelerometry study. Other than the accelerometry data, a rich set of biomedical information was also collected on the study participants, such as health record linkage, self-reported questionnaire and genetic data.

We preprocessed all the datasets by manual quality checks for unrealistic high values for accelerometry (>200 mg), parsing successes, polysomnography alignment, and visual inspection.

### 6. Model development

#### 6.1. Self-supervised pre-training

To obtain a feature extractor by leveraging a large amount of unlabelled data from the UK Biobank, we applied multi-task self-supervised learning following [8]. In self-supervision pre-training, the model was designed to discriminate whether a set of binary transformations have been applied to the signal. We selected reversal, permutation, and time-warping as potential self-supervised learning because they are suitable for learning spatiotemporal patterns.

The feature extractor was built on top of ResNet-17 V2 [9] with 1D convolution, in total, with 10M parameters. Each feature vector is of size 1024. We used cross-entropy as the cost function, with each task having the same weight to balance the features learned from each task. In the training procedure, we applied axis swap and rotation as data augmentation to obtain a representation that is orientation invariant. During training time, we used a batch size of 2000 as a larger batch size was found to produce features with better quality. Adam [10] was used for optimisation with a learning rate of 1e-3. We distributed the training across 4 Tesla V100-SXM2 GPUs with 32GB. Early-stopping with a patience of five steps was used to avoid overfitting. It took about 420 GPU hours for the model to converge. More details can be found in [8].

#### 6.2. SleepNet training

We used the pre-trained ResNet from self-supervision as the base model for feature extraction. Then, we appended two layers of Bi-directional Long-Short-Term-Memory (LSTM) layers of 1024 units to learn the temporal dependencies of the model [11]. In the end, we had two fully-connected layers of 512 units to generate the sleep stages. The model was trained to discriminate five sleep stages directly (wake, N1, N2, N3 and REM). To obtain the three-class output, we combined NREM I, II, and III into the NREM class. Likewise, we combined NREM I, II, III and NREM into the sleep class to obtain the two-class output.

The learning rate was set to be 1e-3. We also set the gradient clapping to 1 to avoid exploding gradient for LSTM. We used weighted Cross-Entropy as the objective function and weighted each class with the inverse of its frequency to account for the imbalanced dataset. We also used rotation and axis swap to augment the input data to obtain a direction-invariant model. Each training mini-batch consisted of five participants. For each individual, we selected four 1.5-hour sequences with random starting points to avoid overfitting to the study protocol, where the beginning and the end of the sequence are always the “wake” class. The model was trained on a Tesla V100-SXM2 with 32GB of memory. It took about 12 hours for the model to converge. The model performance was reported using five-fold subject-wise cross-validation. We first split the data into train/test with a ratio of 8:2. We further split the train set into train/validation with a ratio of 8:2. We used early stopping with a patience of ten steps to avoid overfitting on the validation set in each cross-validation fold.

**Table 2:** Hand-crafted features.

| Handcrafted features | Notes |
| --- | --- |
| Sleep features [12] |  |
| ENMO | All sleep features have 12 derived variables:<br>mean, std, min, max, entropy 20 bins (low resolution),<br>entropy 200 bins (high resolution), median absolute derivation,<br>and mean difference between neighbouring windows. |
| Angle Z |  |
| Locomotor inactivity during sleep |  |
| Axis features [13] |  |
| Mean | 1 per axis |
| Standard deviation | 1 per axis |
| Range | 1 per axis |
| Inter-quantile-range | 1 per axis |
| Correlation of variations | 1 per axis |
| Features on the vector norm [13] | $\text{norm} = \sqrt{x^2 + y^2 + z^2}$ |
| Mean |  |
| Standard deviation |  |
| Inter-quantile-range |  |
| Median absolute derivation |  |
| Kurtosis |  |
| Skew |  |
| Truncated ENMO |  |
| Absolute value of ENMO |  |
| Entropy |  |
| Dominant Frequency |  |
| Total power |  |
| Dominant frequencies | 3 features: 0.3-5 Hz, 0.3-15 Hz, and 0.6-2.5 Hz |
| Dominant frequency power | 3 features: 0.3-5 Hz, 0.3-15 Hz, and 0.6-2.5 Hz |
| Second dominant frequency | 1 feature: 0.3-15 Hz |
| Fourier transform coefficients | 11 features: 1 Hz - 11 Hz |
| Fourier coefficients | 12 features: 1st - 12th coefficient |

**Table 3:** Model performance metric definitions (TP: true positive; TN: true negative; FP: false positive; FN: false negative)

| Metric | Definition |
| --- | --- |
| Precision | $\frac{TP}{TP+FP}$ |
| Sensitivity/Recall | $\frac{TP}{TP+FN}$ |
| Specificity | $\frac{TN}{TN+FP}$ |
| Accuracy | $\frac{TP+TN}{TP+TN+FP+FN}$ |
| F1 | $2 \times \frac{\text{precision} \cdot \text{recall}}{\text{precision} + \text{recall}}$ |
| Kappa | $1 - \frac{1-p_o}{1-p_e}$<br>$p_o$: relative observed agreement<br>$p_e$: expected agreement probability |
| Balanced accuracy | $\frac{1}{n} \sum_i \text{Accuracy}_{class_i}$ |

**Table 4:** Sleep parameter definitions: total sleep duration (TSD), rapid-eye-movement (REM), non-rapid-eye-movement (NREM), sleep onset latency (SOL), wake after sleep onset (WASO), and sleep efficiency (SE).

| Parameter | Definition |
| --- | --- |
| Total sleep duration (TSD) | The total time spent in sleep during the recording period per day. |
| Overnight sleep duration | The longest sleep window duration (max one hour of sleep discontinuity allowed) over a noon-to-noon interval. |
| Time in bed | The amount of time spent in bed: A person might not be asleep during this period. Our time in bed was estimated using a random forest model that was trained using data from sleep diaries. |
| Sleep onset latency (SOL) | The time difference between when one gets in bed and the sleep onset. The sleep onset (SOL) is defined as the first occurrence of three consecutive 30-sec sleep windows. |
| Wake after sleep onset (WASO) | The amount of wake time spent after the sleep onset during the longest sleep window. |
| Sleep efficiency (SE) | SE for sleep window after device-detected sleep onset: $\frac{\text{Overnight sleep duration}}{\text{time in bed}}$ |
| REM duration | The total time spent in the REM stage. |
| REM ratio | $\frac{\text{REM duration}}{\text{TSD}}$ |
| NREM duration | The total time spent in the NREM I, II, and III stages. |
| NREM ratio | $\frac{\text{NREM duration}}{\text{TSD}}$ |

### 7. UK Biobank analysis

**Table 5:** Code table for UK Biobank variables used in the study.

| Variable | Code name |
| --- | --- |
| Month of birth | p52 |
| Year of birth | p34 |
| Device wear time | p90010 |
| Sex | p31 |
| Ethnicity | p21000 |
| Smoking status | p20116 |
| Alcohol consumption | p1558 |
| Education qualification | p6138 |
| Body mass index | p21001 |
| Employment status | p6142 |
| Overall health rating | p2178 |
| Self-reported total sleep duration | p1160 |
| Townsend Deprivation Index | p189 |
| Overall accelerometry average | p90012 |
| Self-reported trouble falling/ staying asleep | p1200 |

The UK Biobank variable codes are shown in Table 5. We used the month of birth (p52) and year of birth (p34) along with device wear time (p90010) to compute the age at wear time. Participants were asked about their insomnia symptoms history (p1200) by “Do you have trouble falling asleep at night or do you wake up in the middle of the night?”. Four responses were possible: “never/rarely”, “sometimes”, “usually”, and “prefer not to answer”.

#### 7.1. Sleep and all-cause mortality

The relationship between machine learning-derived sleep architecture estimates and all-cause mortality was assessed using association analyses. The main analysis split the participants into six groups stratified by sleep efficiency cut-off with clinical relevance. Then, five groups were created based on exact hour cut-offs in line with sleep recommendation guidelines for overnight sleep duration [14]. Four groups were created based on percentage cut-offs of clinical relevance for sleep efficiency [15]. In the sensitivity analysis, seven sleep groups were created on exact hour cut-offs to capture the variations in participants with lower and higher sleep durations.

Mortality was determined using death registry data (obtained by UK Biobank from NHS Digital for participants in England and Wales and from the NHS Central Register, National Records of Scotland, for participants in Scotland). For survival analyses, participants were censored at the earliest of UK Biobank’s record censoring date for mortality data (2021-09-30 for participants in England and Wales and 2021-10-31 for participants in Scotland, with country assigned based on baseline assessment centre) and a record of loss to linked health record follow-up (field 191; 2 participants only).

In addition to the exclusions described for the analyses above, for prospective analyses for incident mortality we further excluded the participants if they had a prior hospitalisation for restless syndrome, any cardiovascular disease or cancer (a hospital episode with primary diagnosis G473, I00-I99 or C00-C99).

Models used age as the timescale, and the main analysis was adjusted for sex (male/female), ethnicity (white/non-white), Townsend Deprivation Index of baseline address (split by quarter in the study population), educational qualifications (school leaver, further education, higher education), smoking status (never smoker, exsmoker, current smoker), alcohol consumption (never, <3 times/week, 3+ times/week), and overall activity (measured in milli-gravity units). An additional analysis further adjusted for BMI (categorised as <18.5 kg/m2, 18.5-24.9 kg/m2, 25.0-29.9 kg/m2, 30+ kg/m2). See Supplementary Table 5 for UK Biobank fields).

Results are presented with their 95% confidence intervals. The Floating Absolute Risk approach was used to calculate confidence intervals for the estimate in each group, without contrast to a reference group [16, 17, 18].

In statistical testing using the Grambsch-Therneau test with the Kaplan-Meier transformation, there was some evidence that the joint associations of overnight sleep duration and sleep efficiency with incident mortality violated the proportional hazards assumption (with age as the timescale). However, assessing associations at younger (< 65 years) and older (≥ 65 years) ages did not suggest substantially differing associations by age, and so the overall hazard ratios are presented.

#### 7.2. Reliability assessment for device wear time exclusion criterion

**Figure 2:**
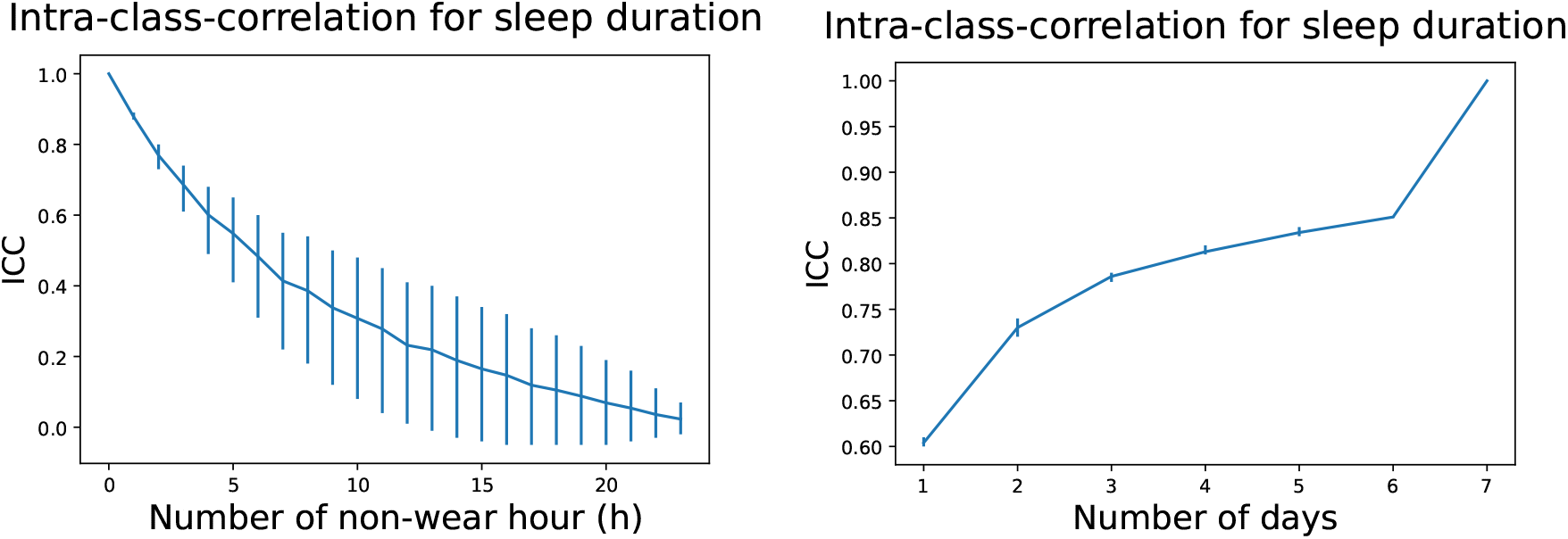
How the intraclass correlation coefficient (ICC) changes with respect to the non-wear hours (h) (left) and the number of wear days (right) in a reliability simulation using data from 27,870 participants that had zero non-wear time across a seven-day period. Mean and 95% confidence intervals are plotted.

We needed to discard participants with too much non-wear time to obtain a stable sleep duration estimate. Ideally, all the participants would have perfect seven-day device wear, which was not the case. Thus, we needed to determine the minimum wear time for seven days so that there is a high agreement between sleep duration computed for participants with perfect data and those computed for participants with missing data. To do this, we first selected a subset of 27,870 participants who did not have any non-wear time during the seven-day window. Then, we simulated the missing data by randomly removing one hour from each day or one whole day of data from each week from their recordings. We increased the amount of simulated missing data step-wise until all the data was removed. Then, we compared weekly mean sleep durations computed on data before and after removing the simulated missing periods.

We used the intraclass correlation coefficient (ICC) to determine the acceptable missing time threshold. We selected two-way random-effects, single rater with an absolute agreement, ICC2, to reflect the reliability of our sleep duration measurement if we have missing data in the measurements [19]. Supplementary Figure 2 depicts the ICC mean and 95% confidence intervals for the missing non-wear hour (Supplementary Figure 2 Left)and missing days (Supplementary Figure 2 Right). We used an ICC of 0.75 threshold when deciding the acceptable device wear range. According to the 0.75 cut-off, a maximum of two non-wear hours per day and a minimum of three days per week are suitable for obtaining stable measurements of sleep duration.

**Figure 3:**
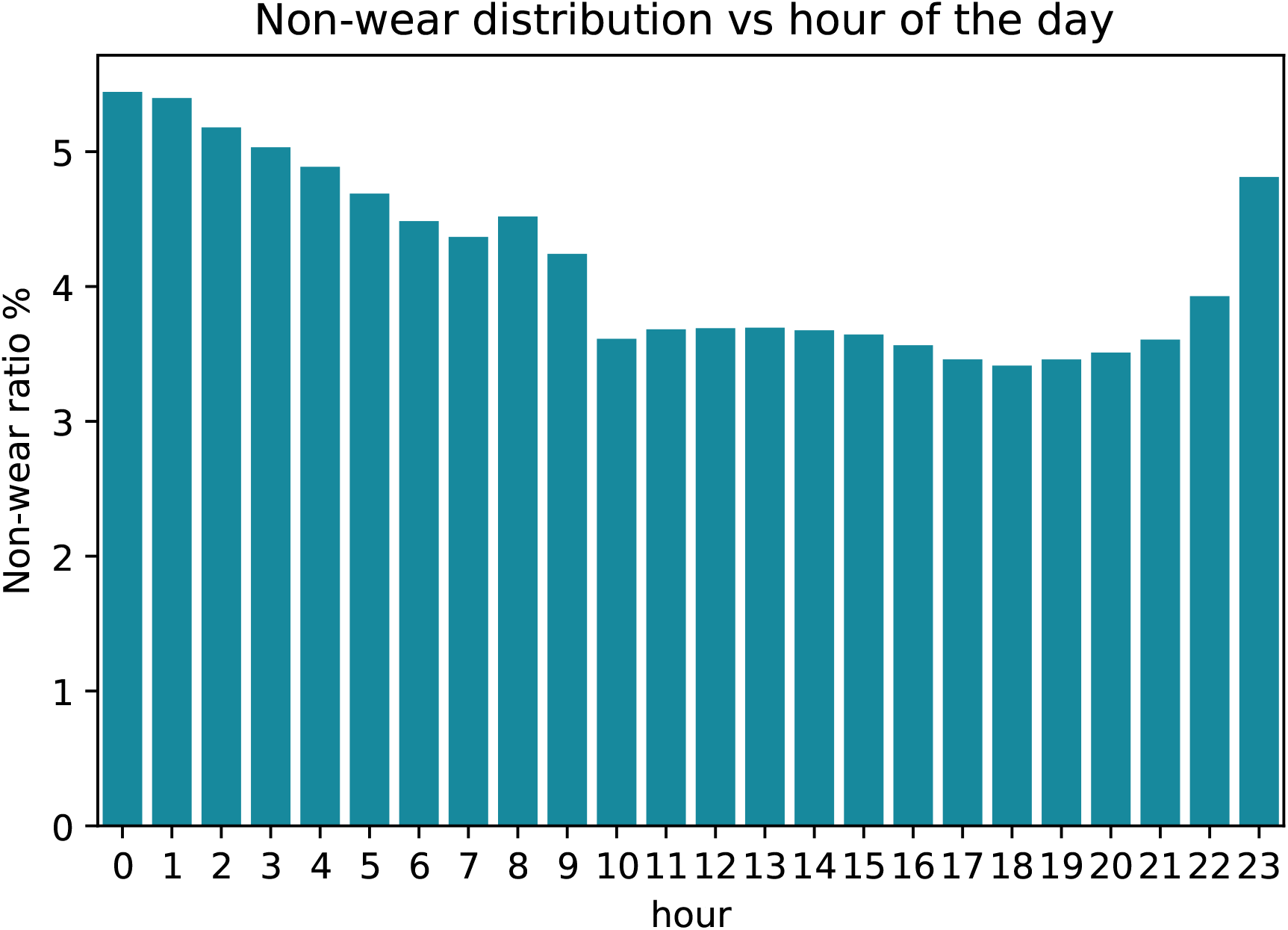
The distribution of non-wear time for all the participants from the UK Biobank.

### 8. Additional Results

#### 8.1. Model performance

**Table 6:** Subject-wise sleep stage classification for benchmark models using internal validation datasets with the Raine Study and the Newcastle cohort: The random forest model was trained using hand-crafted features. SleepNet is the deep recurrent network without pre-training. SleepNet-SSL is the network pre-trained using self-supervision. Five-fold subject-wise performance metrics (mean ± SD) are reported using the internal validation data. REM: rapid-eye-movement sleep, NREM: non-rapid-eye-movement sleep, Kappa score: κ.

| Model | Sleep versus Wake |  |  | Wake versus REM versus NREM |  |  |
| --- | --- | --- | --- | --- | --- | --- |
| | $\kappa$ | Accuracy | F1 | $\kappa$ | Accuracy | F1 |
| Random forest<br>[13, 12] | 0.472±0.192 | 0.756±0.102 | 0.729±0.104 | 0.290±0.149 | 0.507±0.072 | 0.464±0.072 |
| SleepNet | 0.468±0.193 | 0.757±0.103 | 0.727±0.105 | 0.313±0.162 | 0.576±0.112 | 0.535±0.106 |
| SleepNet-SSL | **0.511±0.196** | **0.775±0.105** | **0.750±0.107** | **0.375±0.163** | **0.625±0.116** | **0.573±0.116** |

Supplementary Table 6 shows the model performance comparison between the random forest model that used hand-crafted features and our proposed SleepNet on the internal validation. SleepNet pre-trained with self-supervision had the best performance in both the two-class (κ = 0.511 ± 0.196) and three-class settings (κ = 0.375 ± 0.163). In addition, the area under the receiver operating characteristic curve for the best SleepNet model is0.88 for the two-class setting and0.81 for the three-class setting (Supplementary Figure 4).

**Table 7:** Subject-wise performance sleep classification validation using our best-performing model: All the performance is reported within period in bed. Cohort-specific and pooled performance (Kappa (κ), balanced accuracy, and F1) are shown for both internal and external validation. The pooled performance is calculated by combining all the participants from different datasets. REM: rapid-eye-movement sleep; NREM: non-rapid-eye-movement sleep.

| Dataset | Sleep versus Wake |  |  | Wake versus REM versus NREM |  |  |
| --- | --- | --- | --- | --- | --- | --- |
| | $\kappa$ | Accuracy | F1 | $\kappa$ | Accuracy | F1 |
| Internal validation |  |  |  |  |  |  |
| Raine Gen1 | 0.561±0.161 | 0.791±0.091 | 0.775±0.089 | 0.389±0.152 | 0.623±0.108 | 0.586±0.105 |
| Raine Gen2 | 0.437±0.189 | 0.758±0.101 | 0.712±0.100 | 0.344±0.161 | 0.603±0.115 | 0.552±0.108 |
| Newcastle | 0.394±0.189 | 0.715±0.091 | 0.686±0.103 | 0.285±0.151 | 0.513±0.078 | 0.467±0.085 |
| **Pooled internal** | 0.509±0.184 | 0.777±0.097 | 0.748±0.099 | 0.369±0.158 | 0.613±0.112 | 0.571±0.108 |
| External Validation |  |  |  |  |  |  |
| Leicester | 0.278±0.141 | 0.678±0.072 | 0.633±0.075 | 0.253±0.122 | 0.527±0.086 | 0.488±0.082 |
| Pennsylvania | 0.468±0.225 | 0.807±0.117 | 0.725±0.118 | 0.374±0.172 | 0.626±0.092 | 0.565±0.097 |
| **Pooled external** | 0.360±0.205 | 0.734±0.114 | 0.673±0.106 | 0.306±0.157 | 0.570±0.101 | 0.521±0.097 |

**Figure 4:**
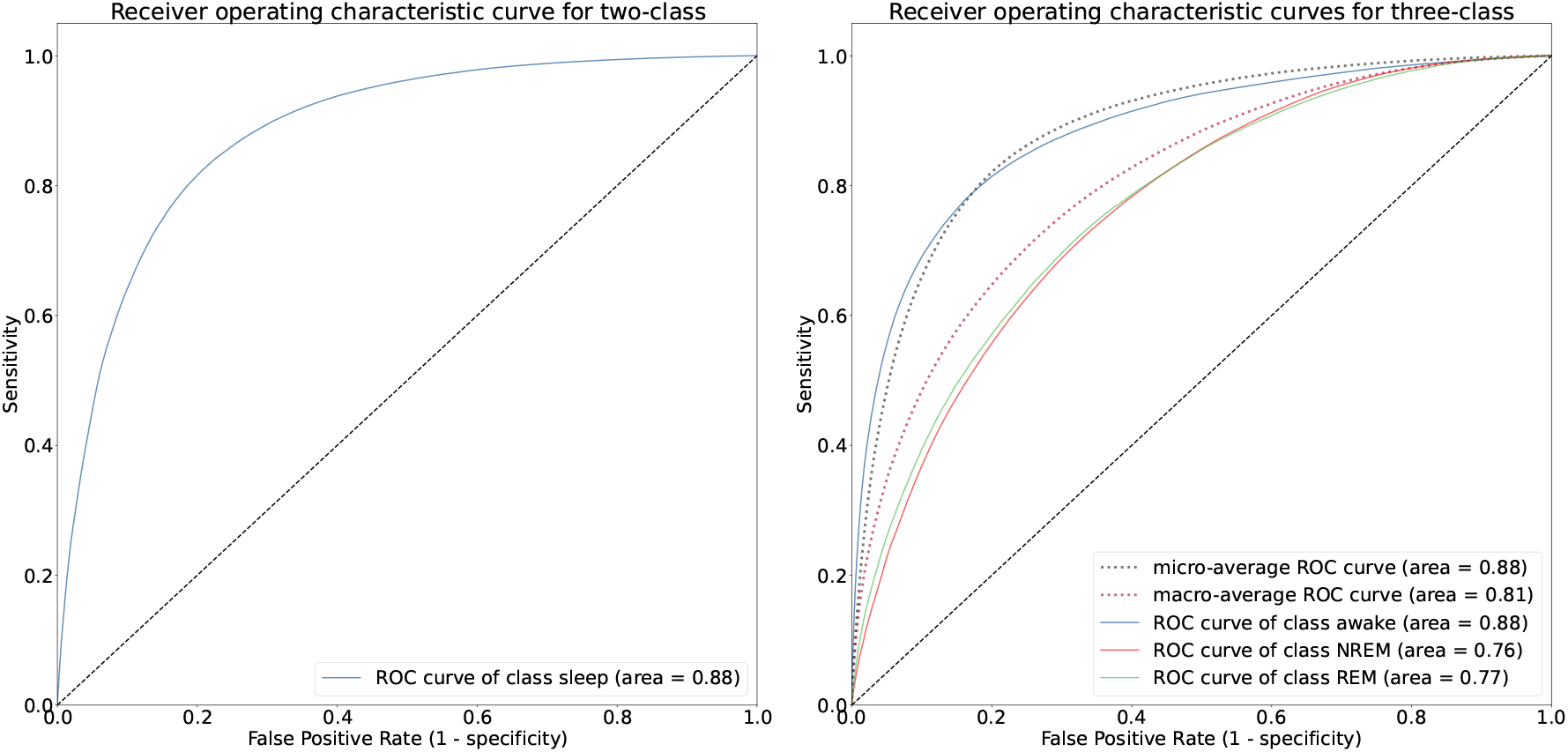
Receiver operating characteristics curves for two-class (wake/sleep) and three-class (wake/REM/NREM) settings on the internal validation dataset using our best performing model self-supervised SleepNet. REM: rapid-eye-movement sleep, NREM: non-rapid-eye-movement sleep.

**Table 8:**
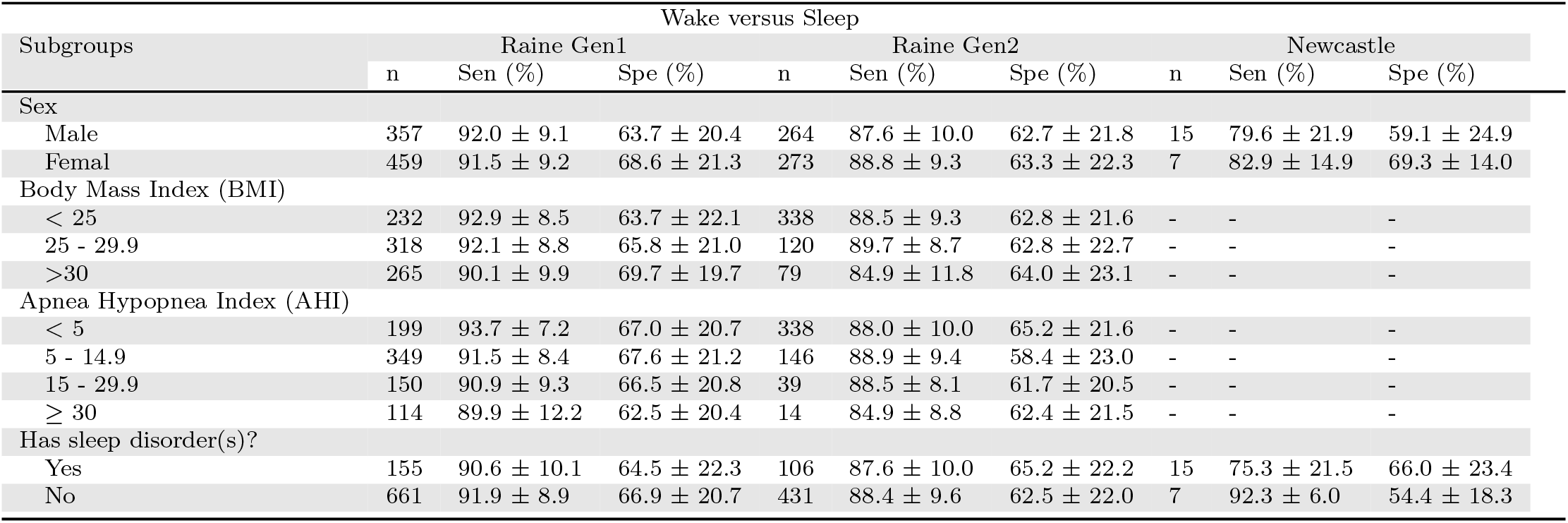
Model characteristics on the internal validation datasets (wake versus sleep): subject-wise performance metrics (mean ± SD) are reported using the internal validation data. Sen: sensitivity, Spe: specificity. Wake is the negative class and the sleep is the positive class when calculating model performance.

**Table 9:**
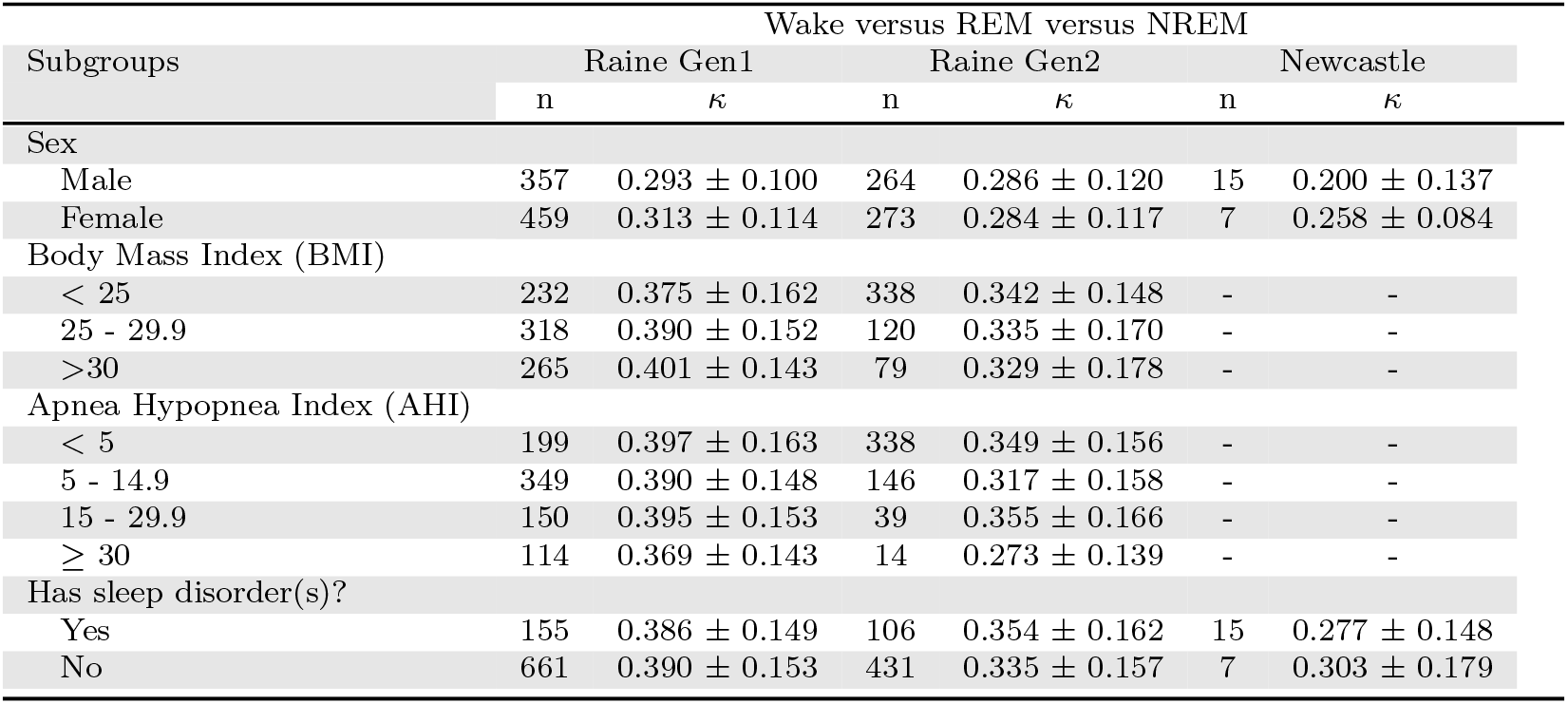
Model characteristics on the internal validation datasets (wake versus REM versus NREM): subject-wise performance metrics (mean ± SD) are reported using the internal validation data. REM: rapid-eye-movement, NREM: non-rapid-eye-movement, Kappa score: κ.

**Table 10:** Model characteristics on the internal validation datasets (wake versus REM versus NREM I, II, III): subject-wise performance metrics (mean ± SD) are reported using the internal validation data. REM: rapid-eye-movement, NREM: non-rapid-eye-movement, Kappa score: κ.

| Subgroups | Wake versus REM versus NREM I, II, III |  |  |  |  |  |
| --- | --- | --- | --- | --- | --- | --- |
|  | Raine Gen1 |  | Raine Gen2 |  | Newcastle |  |
| | n | $\kappa$ | n | $\kappa$ | n | $\kappa$ |
| Sex |  |  |  |  |  |  |
| Male | 357 | $0.279 \pm 0.103$ | 264 | $0.287 \pm 0.120$ | 16 | $0.014 \pm 0.102$ |
| Female | 459 | $0.307 \pm 0.111$ | 273 | $0.285 \pm 0.113$ | 9 | $0.125 \pm 0.106$ |
| Body Mass Index (BMI) |  |  |  |  |  |  |
| < 25 | 232 | $0.295 \pm 0.117$ | 338 | $0.286 \pm 0.110$ | - | - |
| 25 - 29.9 | 318 | $0.309 \pm 0.107$ | 120 | $0.292 \pm 0.127$ | - | - |
| >30 | 265 | $0.307 \pm 0.102$ | 79 | $0.273 \pm 0.140$ | - | - |
| Apnea Hypopnea Index (AHI) |  |  |  |  |  |  |
| < 5 | 199 | $0.307 \pm 0.114$ | 338 | $0.293 \pm 0.116$ | - | - |
| 5 - 14.9 | 349 | $0.309 \pm 0.108$ | 146 | $0.264 \pm 0.118$ | - | - |
| 15 - 29.9 | 150 | $0.309 \pm 0.104$ | 39 | $0.299 \pm 0.127$ | - | - |
| $\geq 30$ | 114 | $0.283 \pm 0.104$ | 14 | $0.274 \pm 0.131$ | - | - |
| Has sleep disorder(s)? |  |  |  |  |  |  |
| Yes | 155 | $0.286 \pm 0.107$ | 106 | $0.297 \pm 0.131$ | 15 | $0.213 \pm 0.136$ |
| No | 661 | $0.309 \pm 0.108$ | 431 | $0.283 \pm 0.115$ | 7 | $0.230 \pm 0.099$ |

#### 8.2. Cohort-specific performance against polysomnography using SleepNet

**Figure 5:**
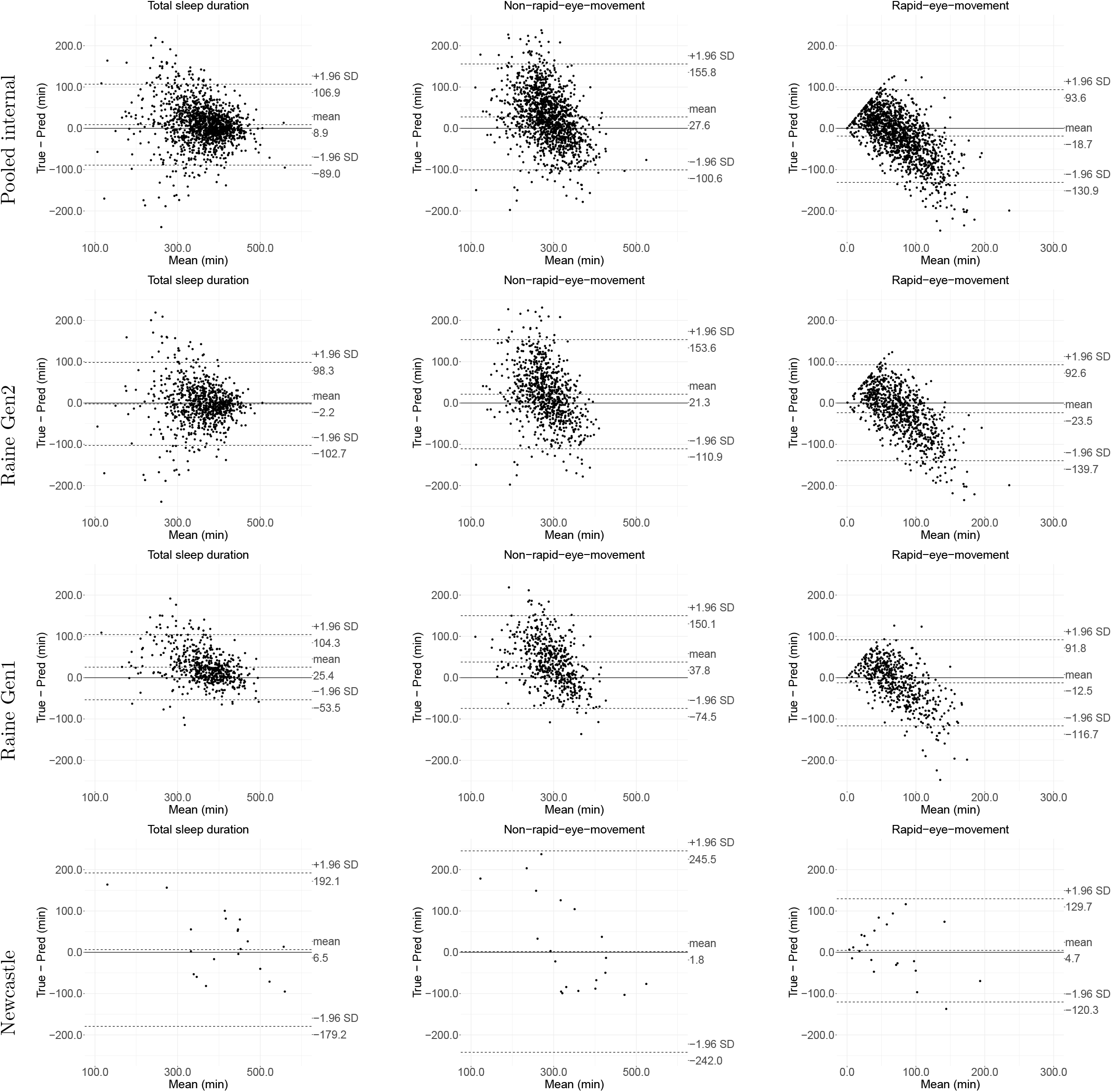
Agreement assessment via Bland-Altman plots for internal validation: total sleep duration (TSD), non-rapid-eye-movement sleep (NREM), and rapid-eye-movement sleep (REM).

**Figure 6:**
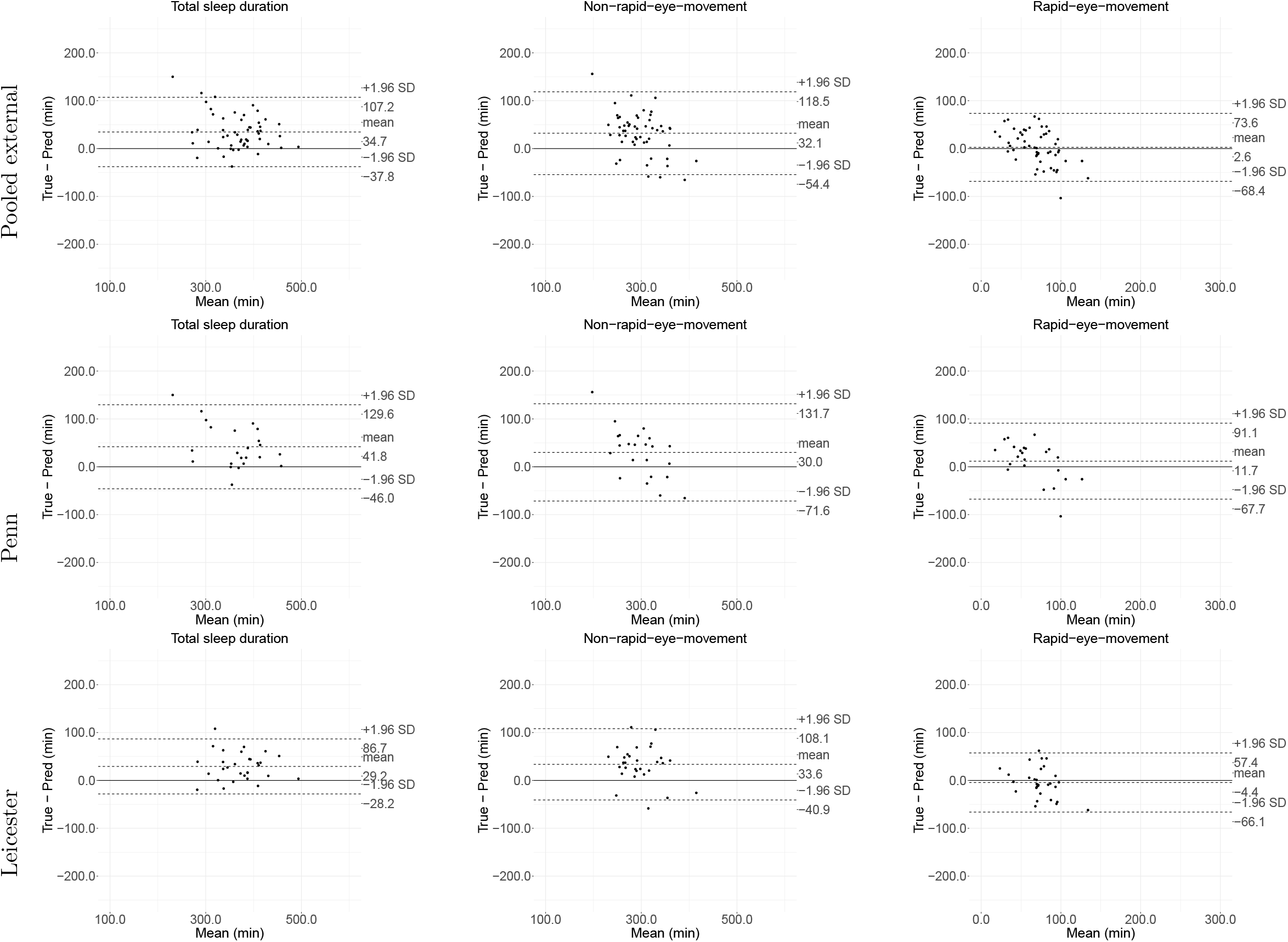
Agreement assessment via Bland-Altman plots for external validation: total sleep duration, wake after sleep onset (WASO), non-rapid-eye-movement sleep (NREM), and rapid-eye-movement sleep (REM).

**Figure 7:**
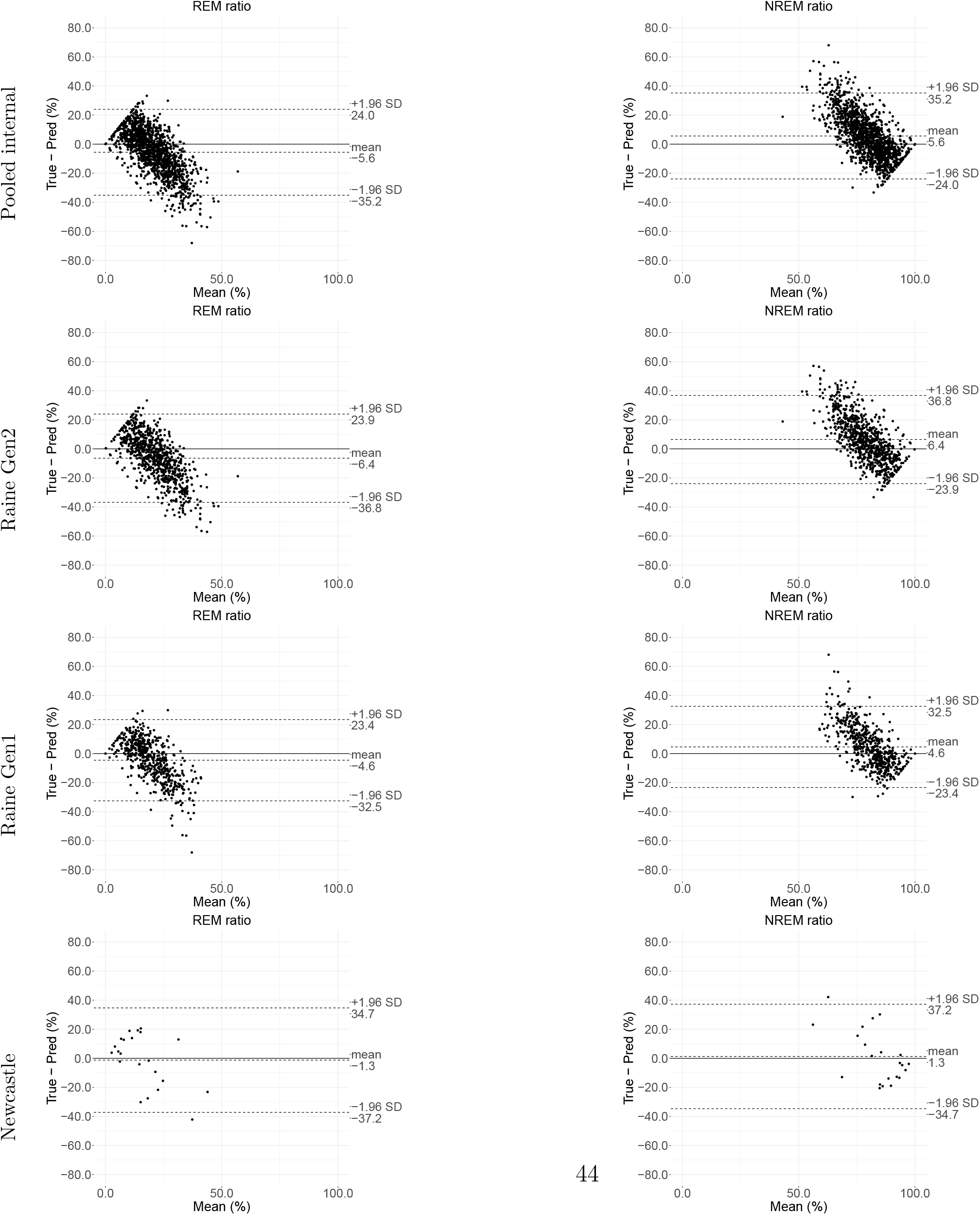
Agreement assessment via Bland-Altman plots for internal validation: non-rapid-eye-movement sleep (NREM) ratio, and rapid-eye-movement sleep (REM) ratio.

**Figure 8:**
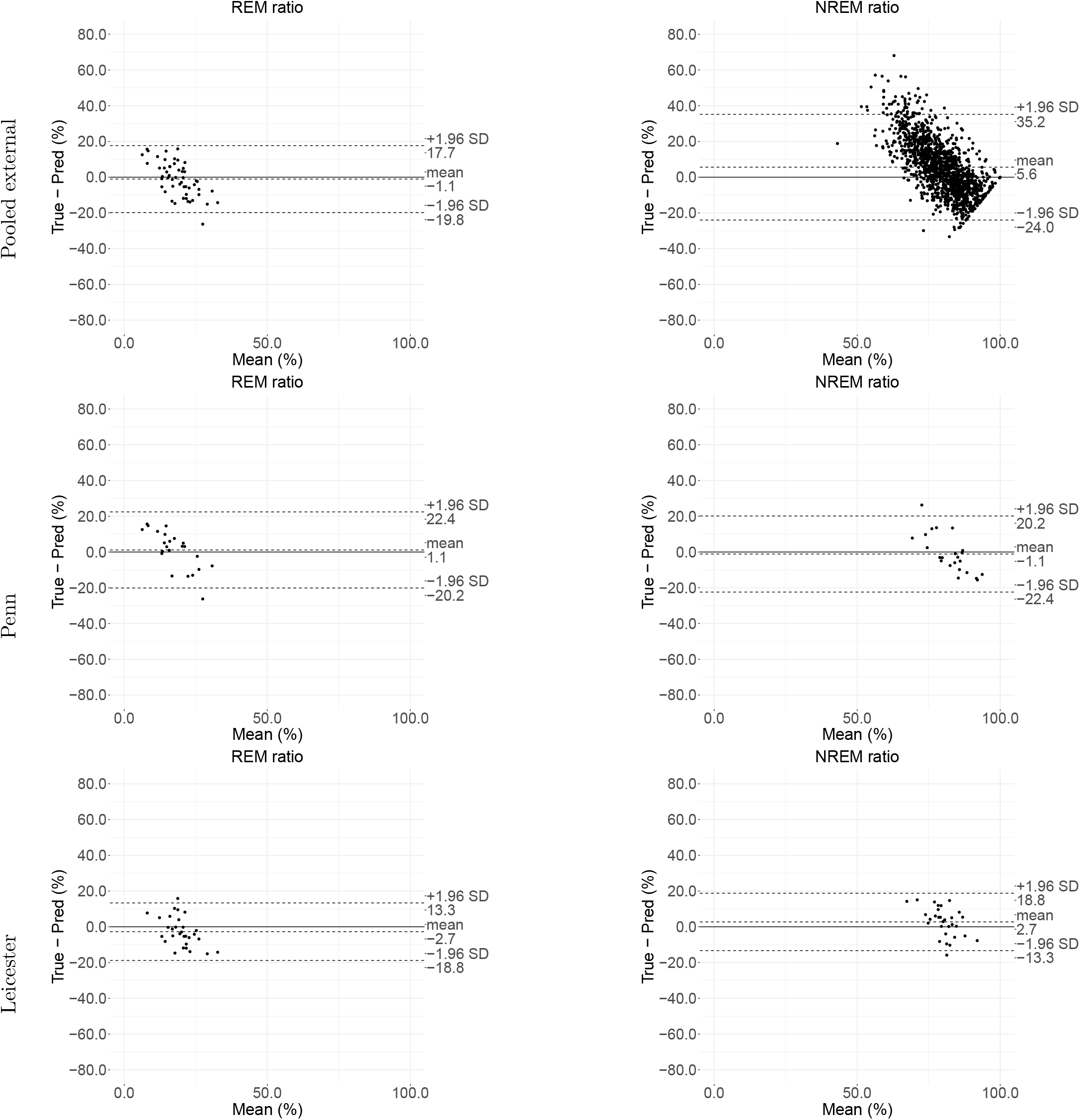
Agreement assessment via Bland-Altman plots for external validation: non-rapid-eye-movement sleep (NREM) ratio, and rapid-eye-movement sleep (REM) ratio.

**Figure 9:**
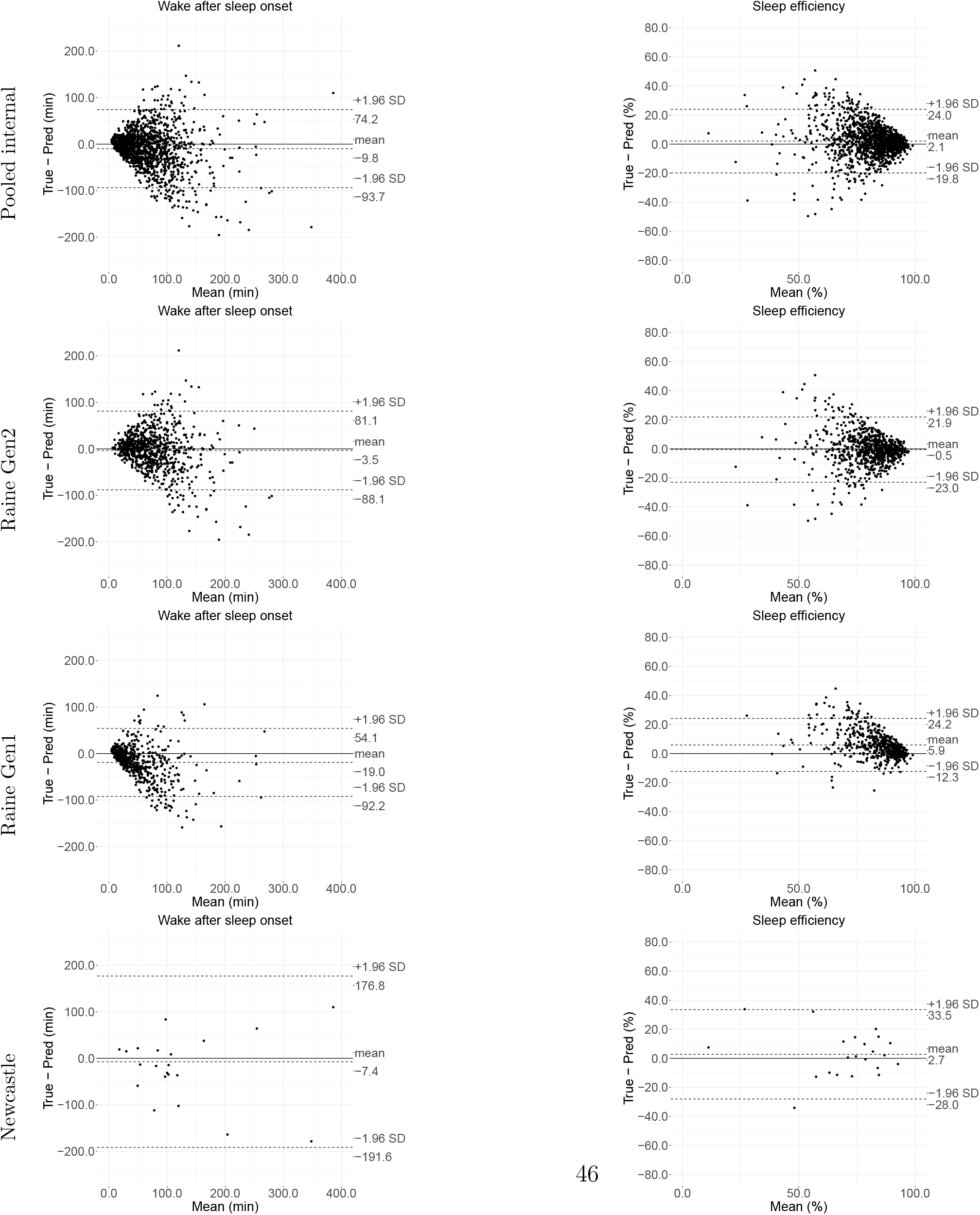
Agreement assessment via Bland-Altman plots for internal validation: wake after sleep onset (WASO), and sleep efficiency (SE).

**Figure 10:**
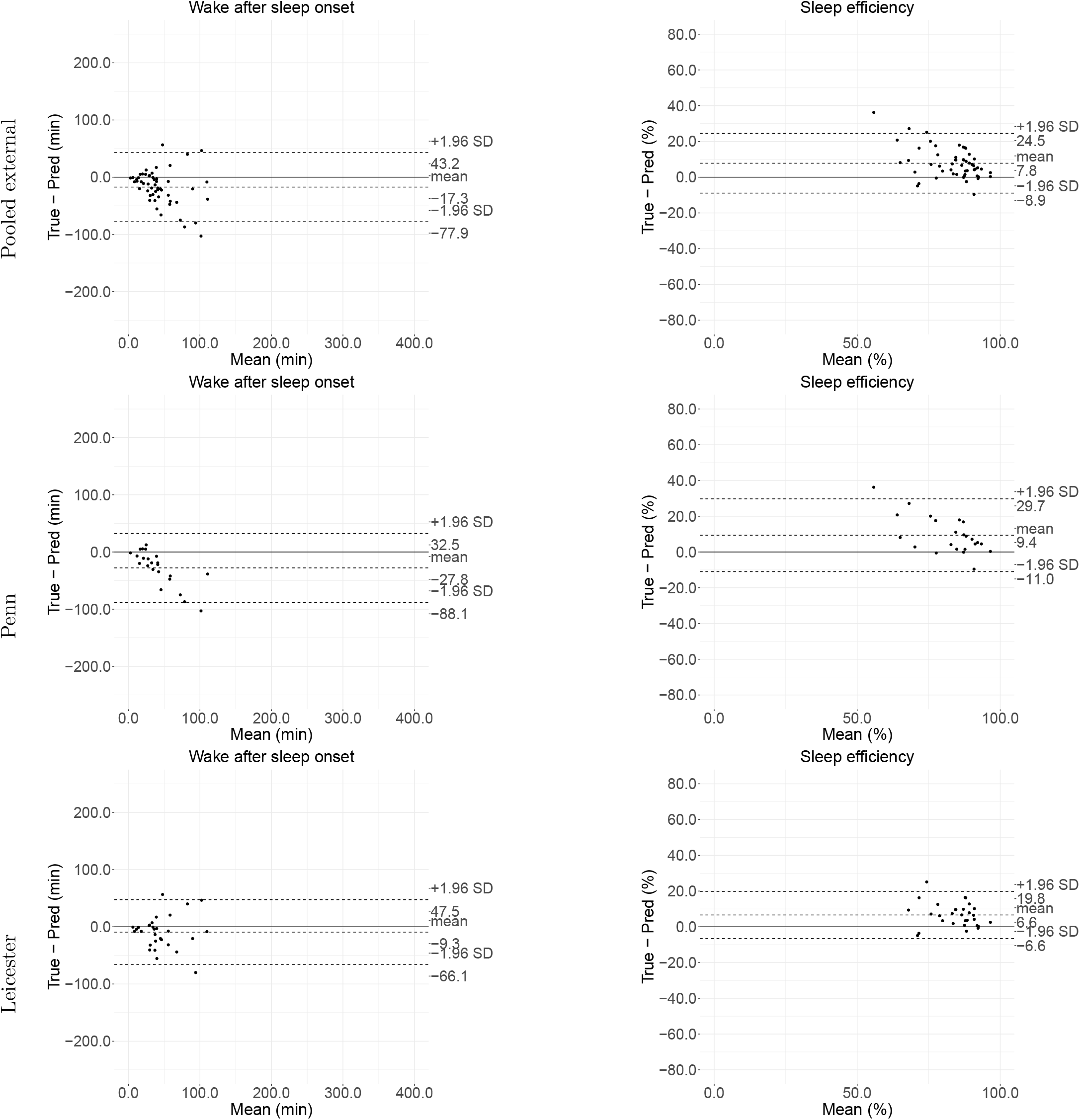
Agreement assessment via Bland-Altman plots for internal validation: wake after sleep onset (WASO), and sleep efficiency (SE).

**Figure 11:**
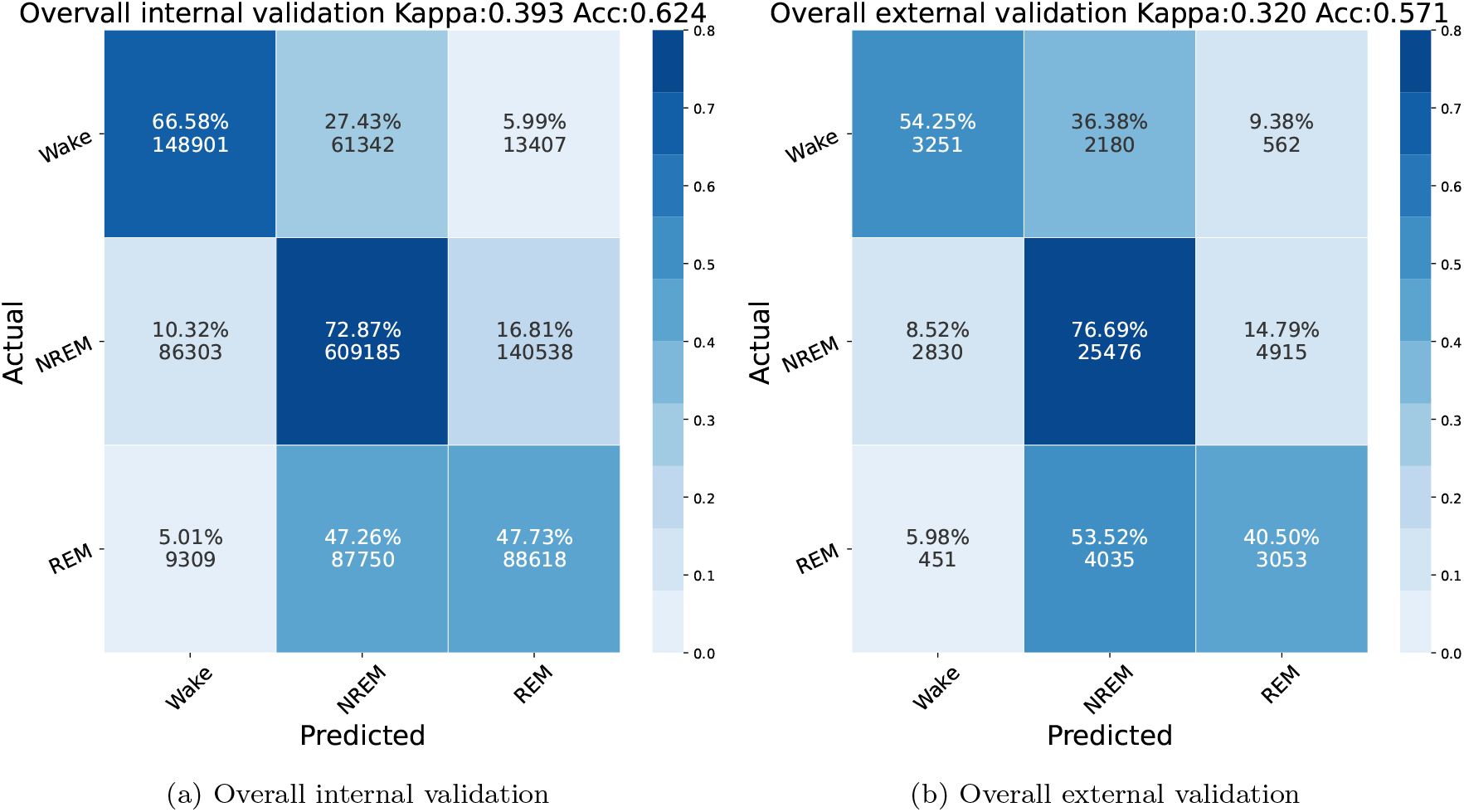
Three class classification (wake/REM/NREM) confusion matrix: epoch-to-epoch Kappa and balanced accuracies are shown. The number of predictions and proportion ratios are shown for each pair of ground-truth and prediction class. REM: rapid-eye-movement sleep; NREM: non-rapid-eye-movement sleep.

**Figure 12:**
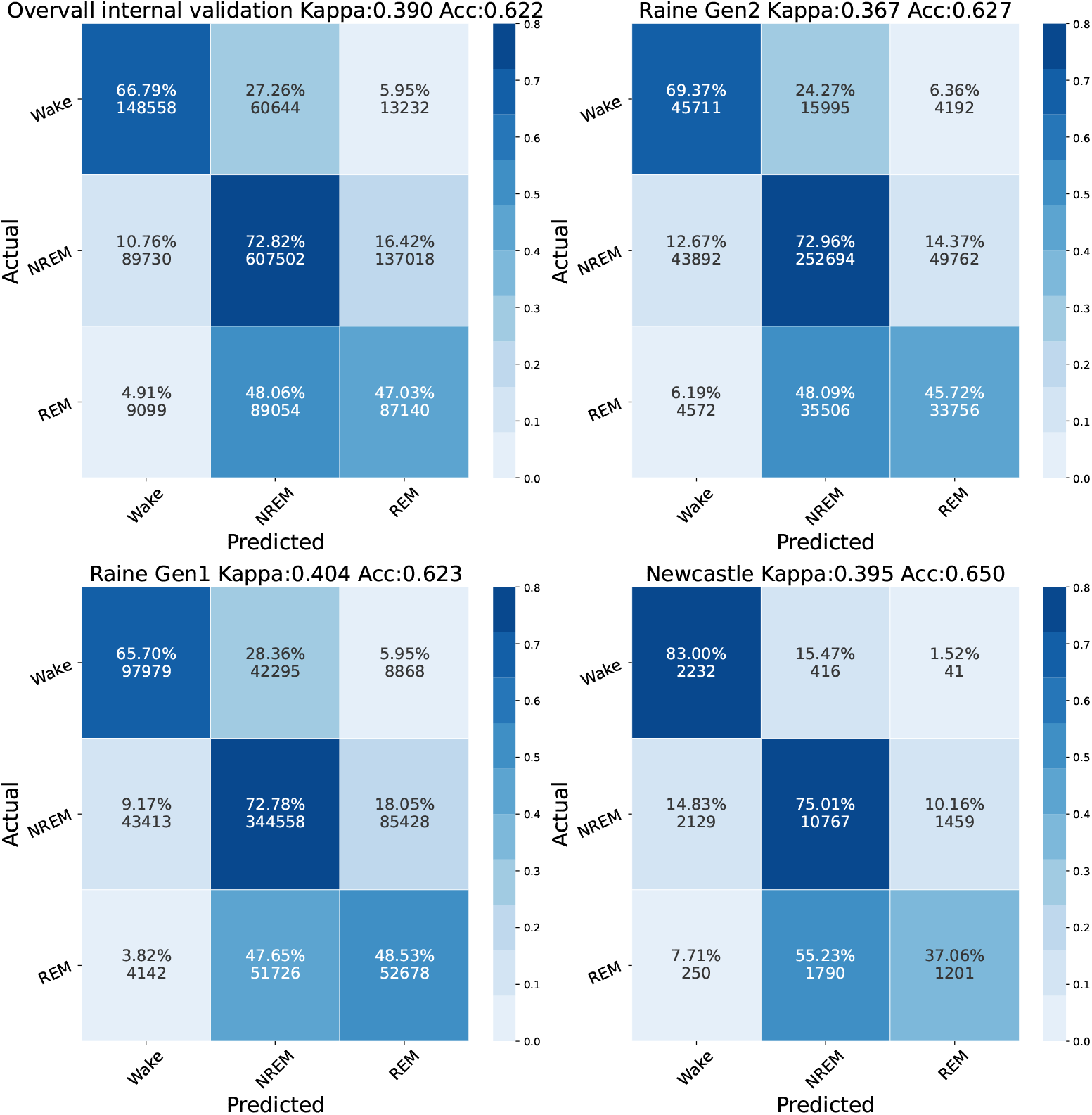
Three-class sleep staging (wake/REM/NREM) for internal validation: epoch-to-epoch Kappa and balanced accuracies are shown. The number of predictions and proportion ratios are shown for each pair of ground-truth and prediction class. REM: rapid-eye-movement sleep; NREM: non-rapid-eye-movement sleep.

**Figure 13:**
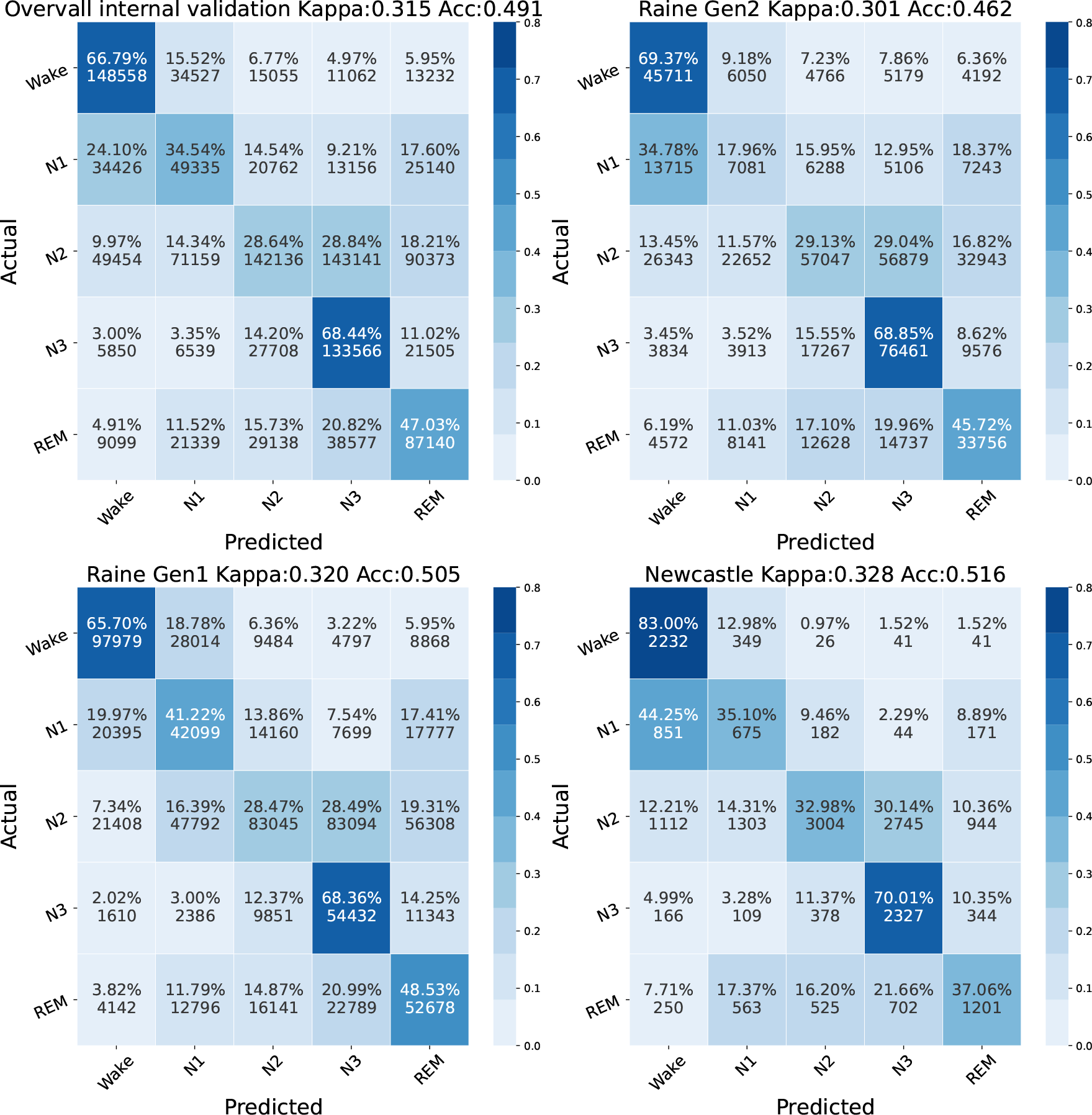
Five-class sleep staging (wake/REM/N1/N2/N3) for internal validation: epoch-to-epoch kappa and balanced accuracies are shown. The number of predictions and proportion ratios are shown for each pair of ground-truth and prediction class. REM: rapid-eye-movement sleep, N1, N2, N3: non-rapid-eye-movement sleep 1, 2, 3.

**Figure 14:**
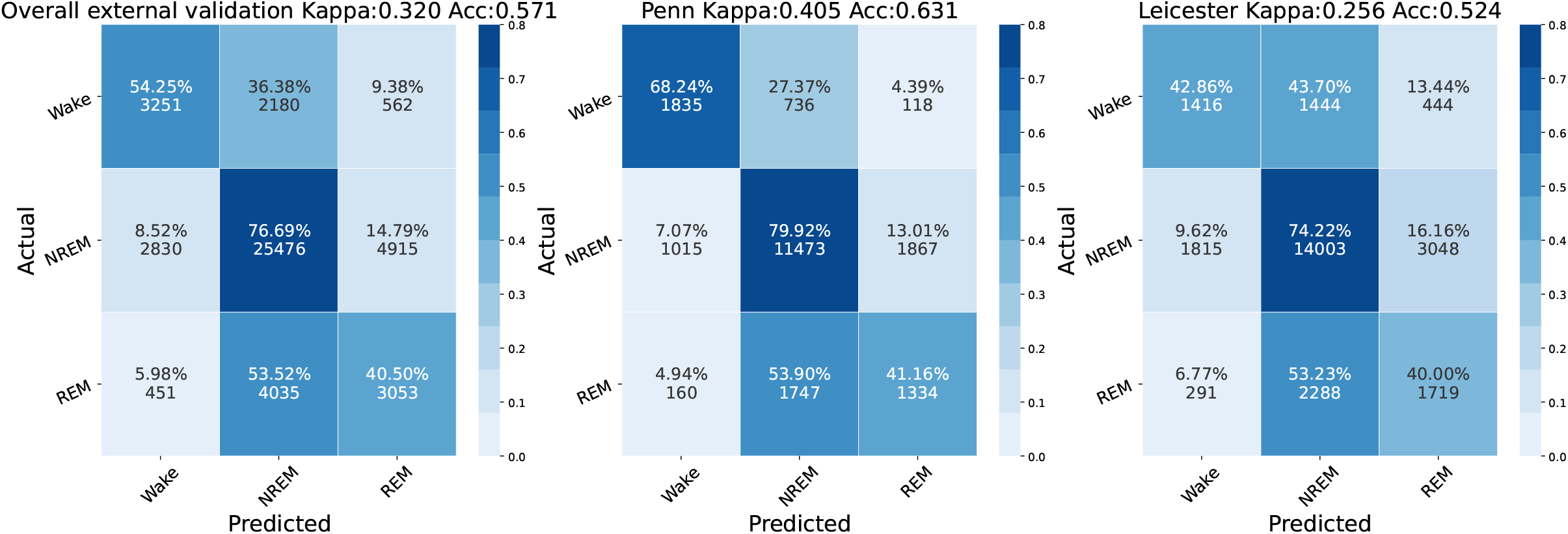
Three-class sleep staging (wake/REM/NREM) for external validation: epoch-to-epoch kappa and balanced accuracies are shown. The number of predictions and proportion ratios are shown for each pair of ground-truth and prediction class. REM: rapid-eye-movement sleep; NREM: non-rapid-eye-movement sleep.

**Figure 15:**
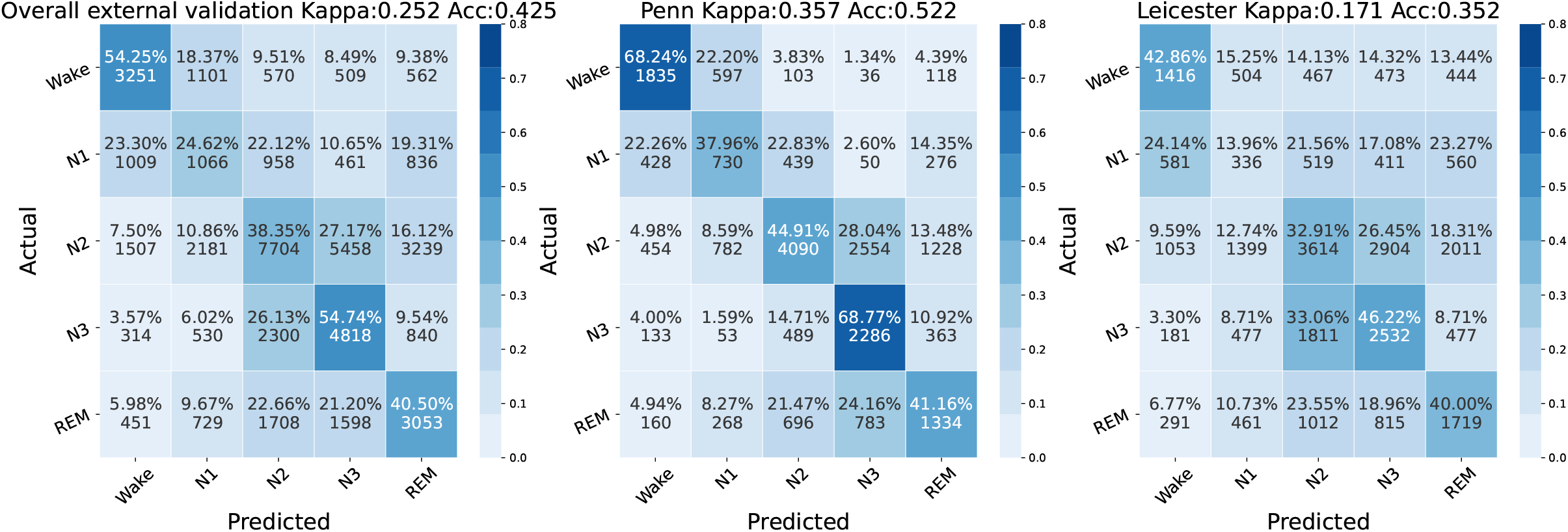
Five-class sleep staging (wake/REM/N1/N2/N3) for external validation: epoch-to-epoch kappa and balanced accuracies are shown. The number of predictions and proportion ratios are shown for each pair of ground-truth and prediction class. REM: rapid-eye-movement sleep, N1, N2, N3: non-rapid-eye-movement sleep 1, 2, 3.

**Figure 16:**
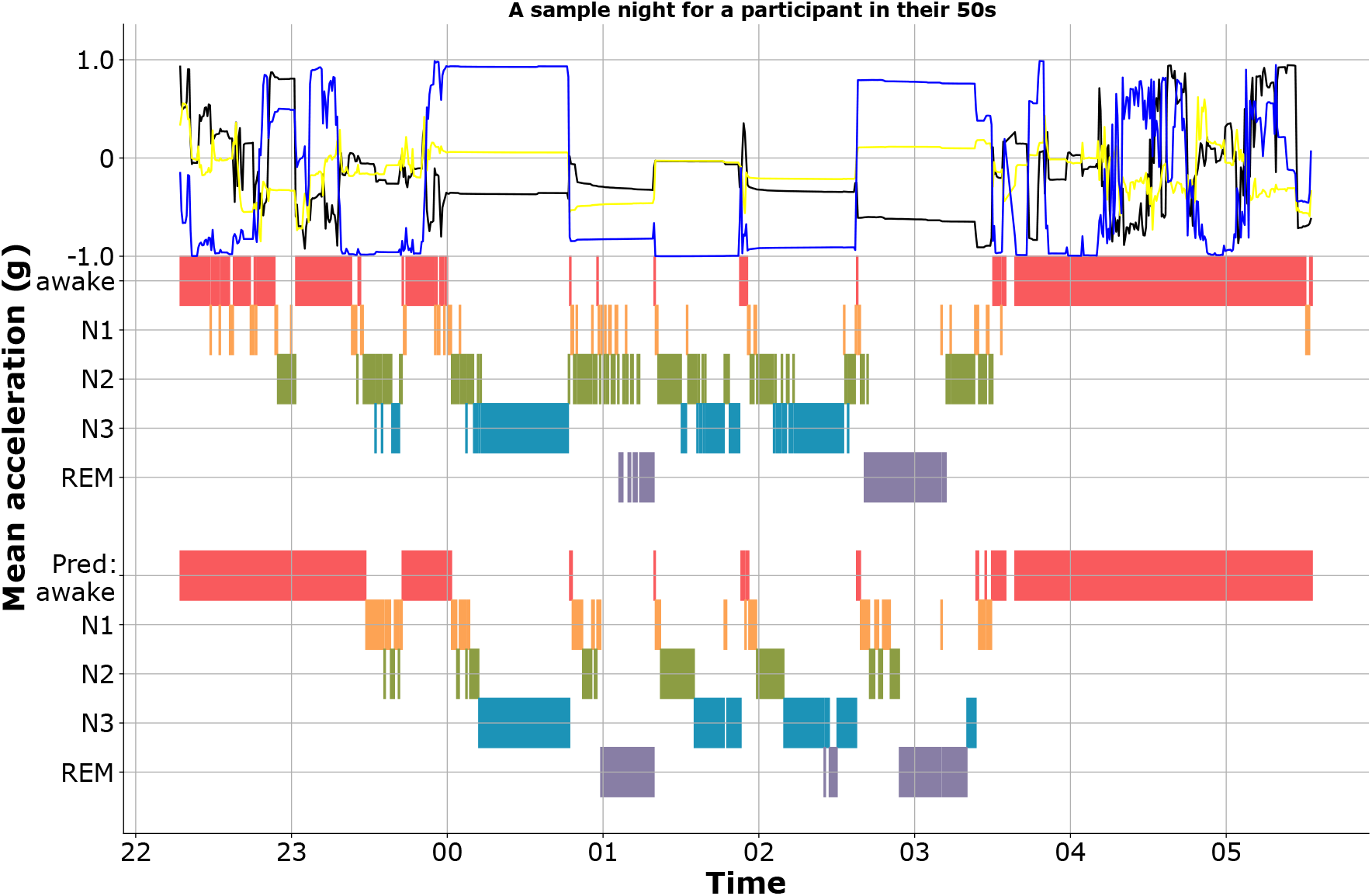
A sample actigram, hypnogram ground truth and prediction for a participant whose sleep stages are well captured: the **top** hypnogram is the ground-truth and the **bottom** hypnogram is the prediction generated by SleepNet based on the actigram. REM: rapid-eye-movement sleep, N1, N2, N3: non-rapid-eye-movement sleep 1, 2, 3.

#### 8.3. Additional results on the sleep variations for the UK Biobank participants

**Figure 17:**
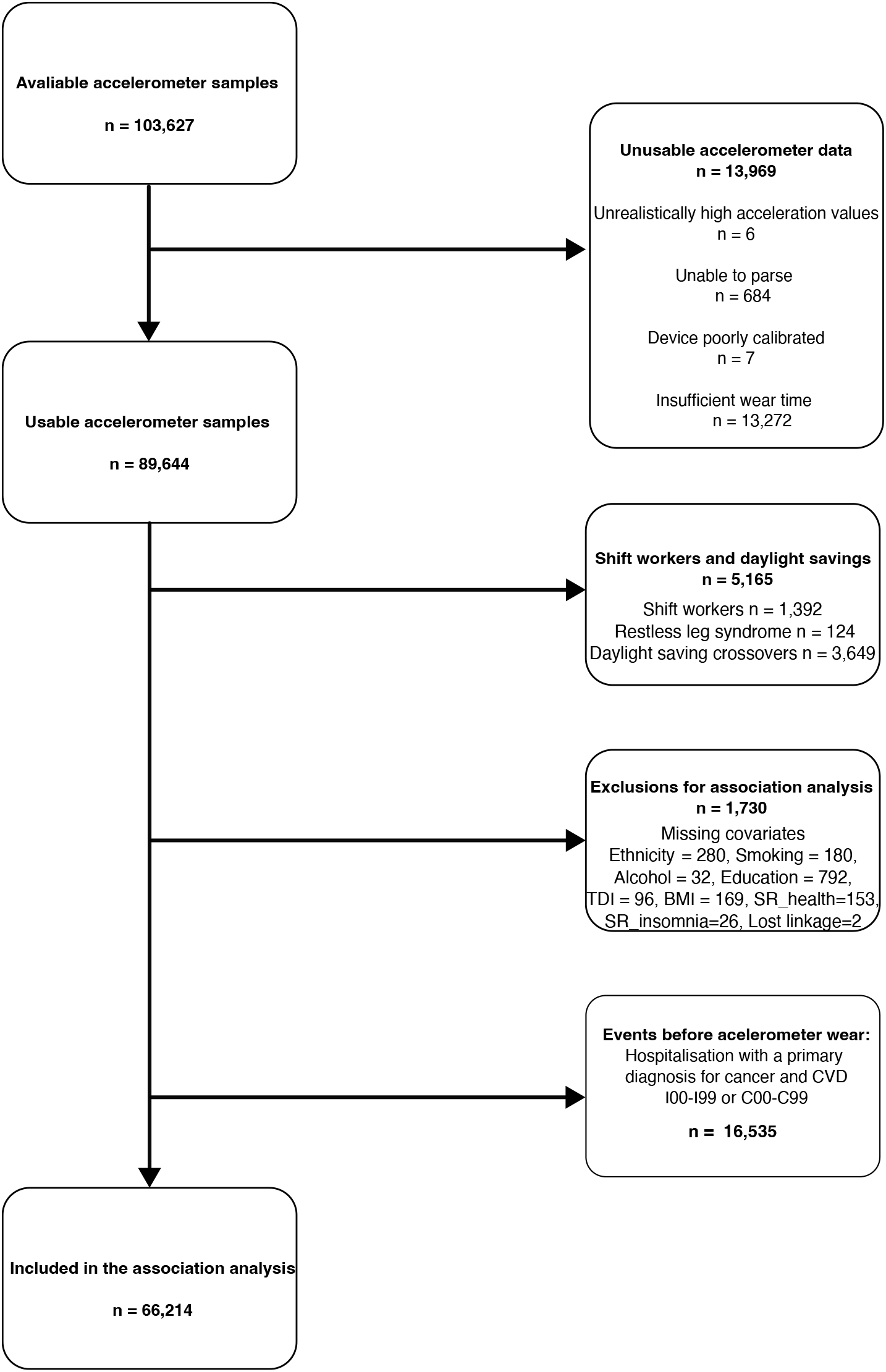
Participant flow diagram for the analysis of sleep and all-cause mortality in the UK Biobank. TDI: Townsend deprivation index, BMI: body mass index, SR health: self-reported overall health, SR insomnia: self-reported insomnia symptoms, CVD: Cardiovascular disease.

**Figure 18:**
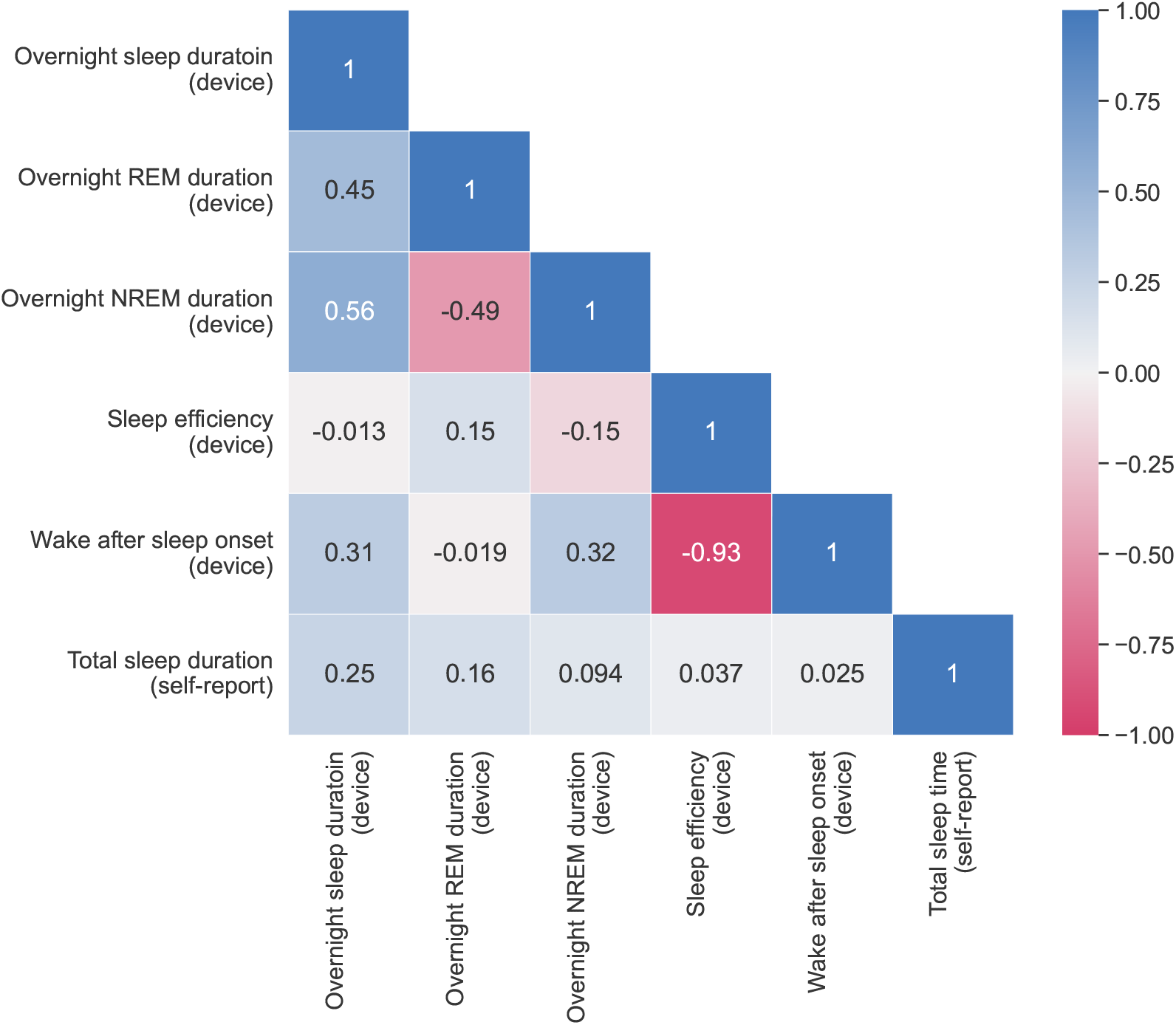
Correlation matrix for device-measured and self-reported sleep parameters on the UK Biobank. The self-reported total sleep duration was obtained via questionnaire at baseline assessment in the UK Biobank. REM: rapid-eye-movement sleep, NREM: non-rapid-eye-movement sleep.

**Figure 19:**
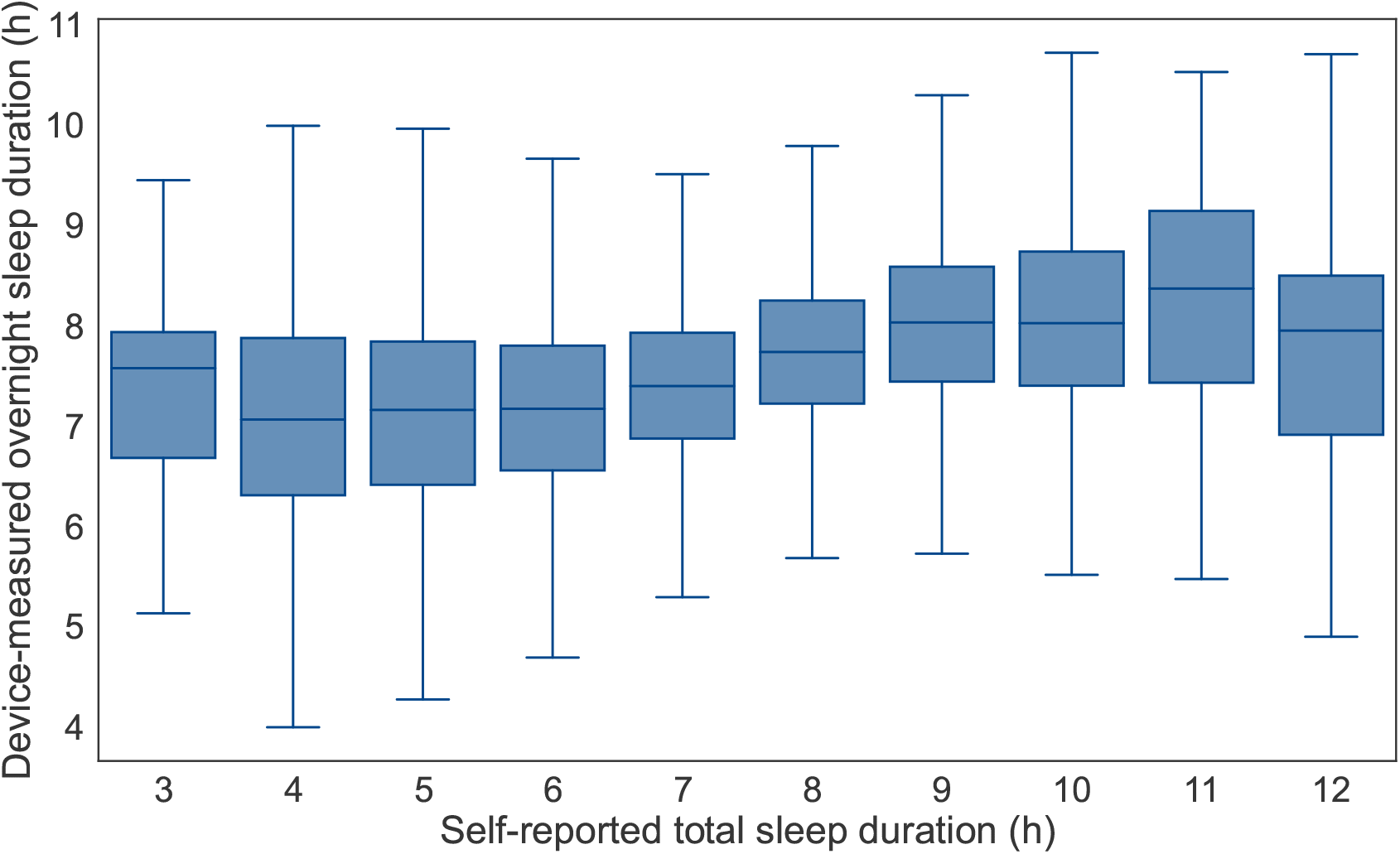
Box plots showing the distributions of device-measured overnight sleep duration against self-reported total sleep duration. The box whiskers reflect the lowest and highest data points that are 1.5 times of the inter-quartile-range from the median.

**Figure 20:**
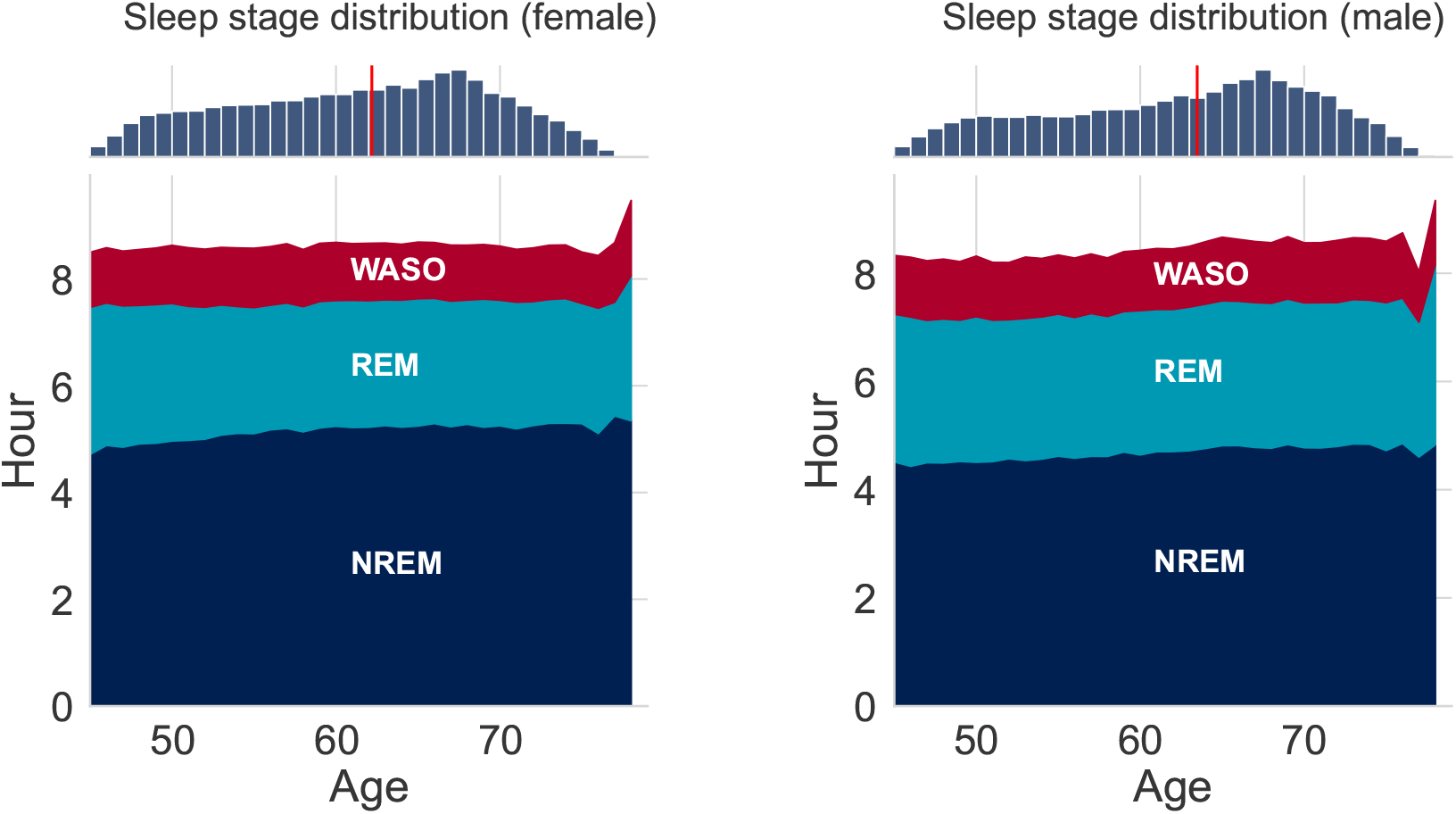
The average device-measured sleep stage distribution with respect to age for both females (left) and males (right) on the UK Biobank. The histograms on the top show the age distribution for the participants. The red vertical line denotes the median age for each sex. WASO: wake after sleep onset; REM: rapid-eye-movement sleep; NREM: non-rapid-eye-movement sleep.

**Figure 21:**
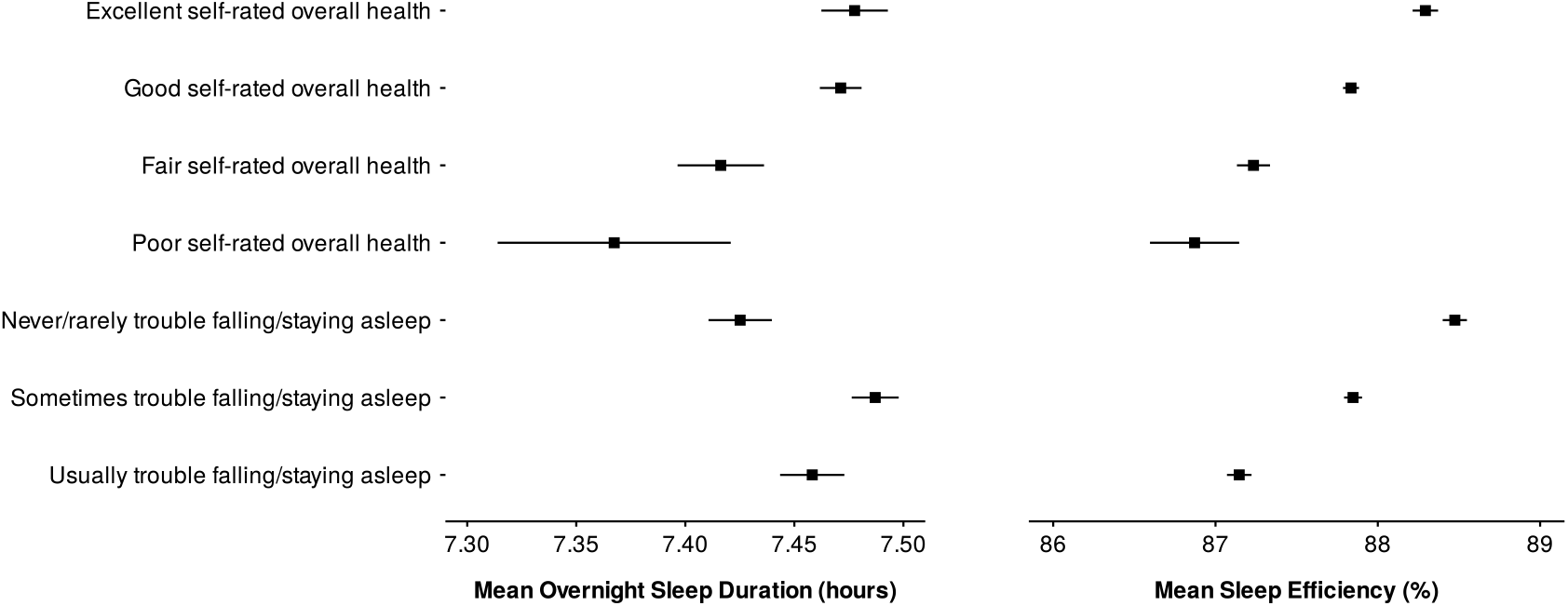
Adjusted marginal mean (95% confidence interval) device-measured mean overnight sleep duration and mean sleep efficiency by self-reported overall health status and insomnia history in the UK Biobank. Mean overnight sleep duration and sleep efficiency were adjusted for age and sex.

**Figure 22:**
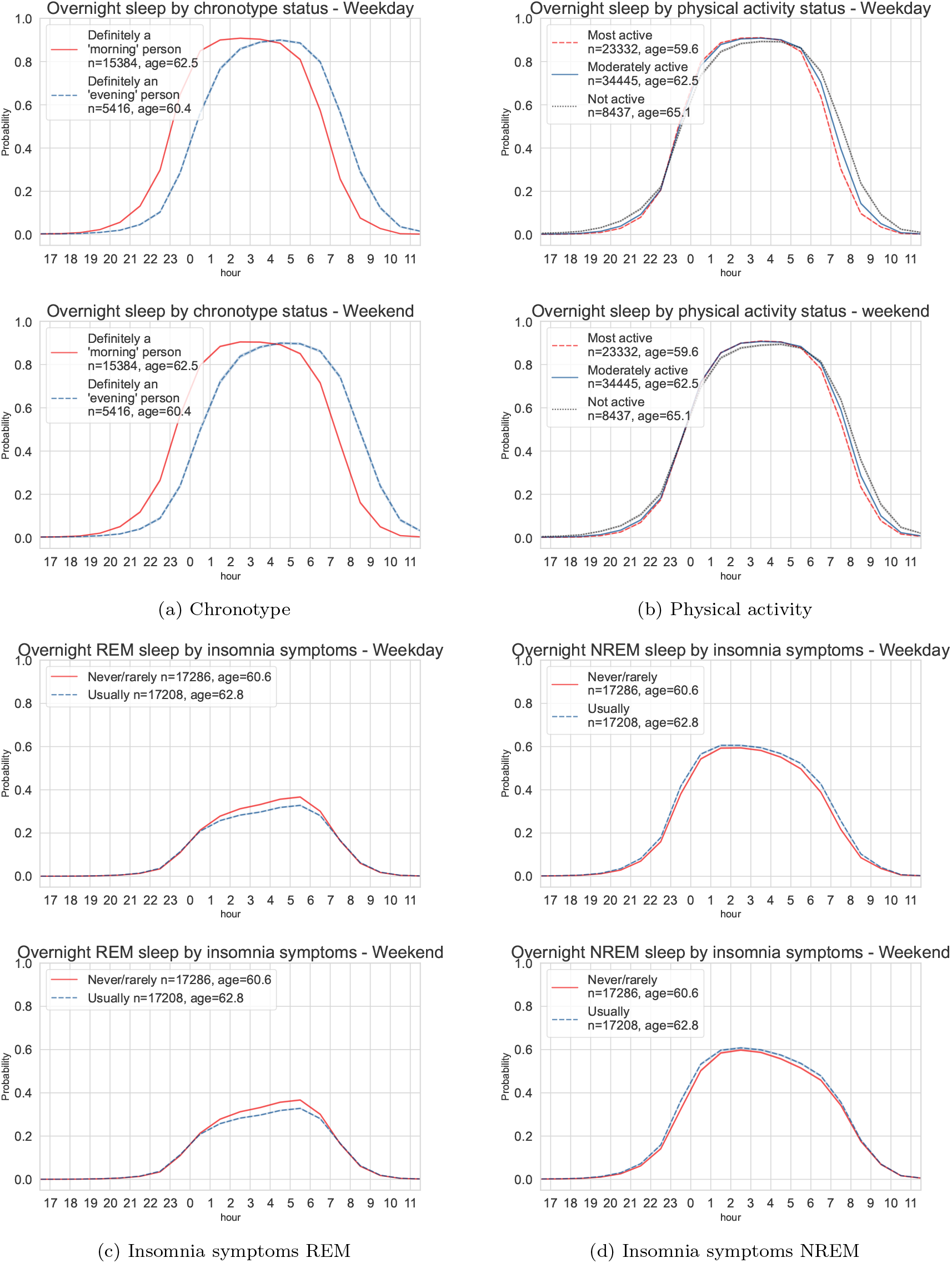
Device-measured sleep probability trajectories throughout the day for the UK Biobank participants (weekday vs weekend). Top: variations of the average overnight sleep probability for the participants with self-reported “morning” and “evening” chronotype (a) and the overnight sleep distributions across thirds of device-measured physical activity level (b). Bottom: variations of the average REM (c) and NREM (d) probability in participants with a history of self-reported insomnia symptoms versus those without. Rapid-eye-movement sleep (REM), and non-rapid-eye-movement sleep (NREM). Areas of squares represent the inverse of the variance of the log risk. And the I bars denote the 95% confidence interval for the floated risks.

**Figure 23:**
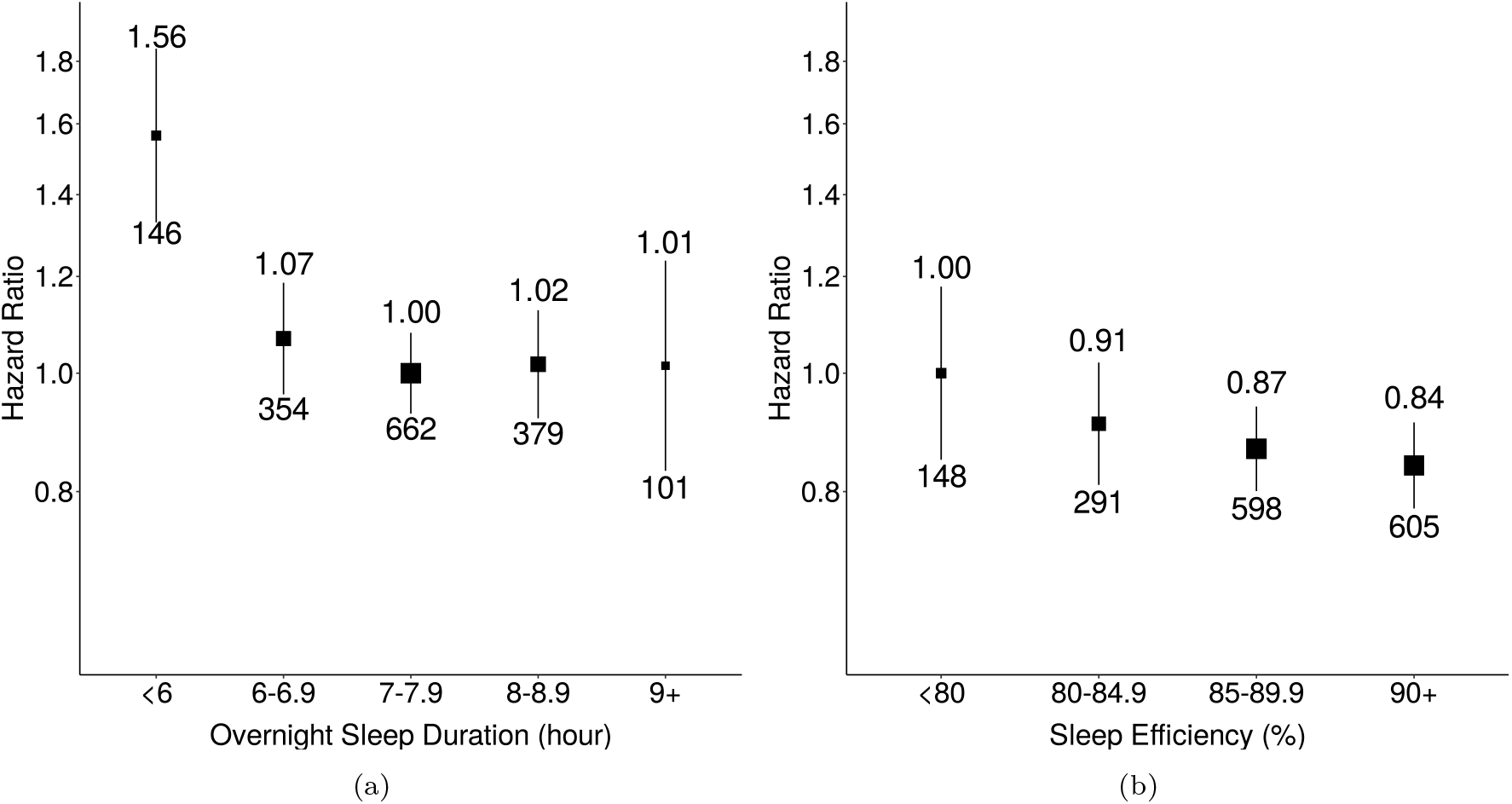
Associations of overnight sleep duration (a) and sleep efficiency (b) with all-cause mortality. The model used 1,642 events among 62,214 participants. We used age as the timescale and adjusted for sex, ethnicity, Townsend Deprivation Index of baseline address (split by quarter in the study population), educational qualifications, smoking status, alcohol consumption (Never, <3 times/week, 3+ times/week), overall activity (measured in milli-gravity units). Areas of squares represent the inverse of the variance of the log risk. The I bars denote the 95% confidence interval for the floated risks.

##### 8.3.1. Models additionally adjusted for body mass index

**Figure 24:**
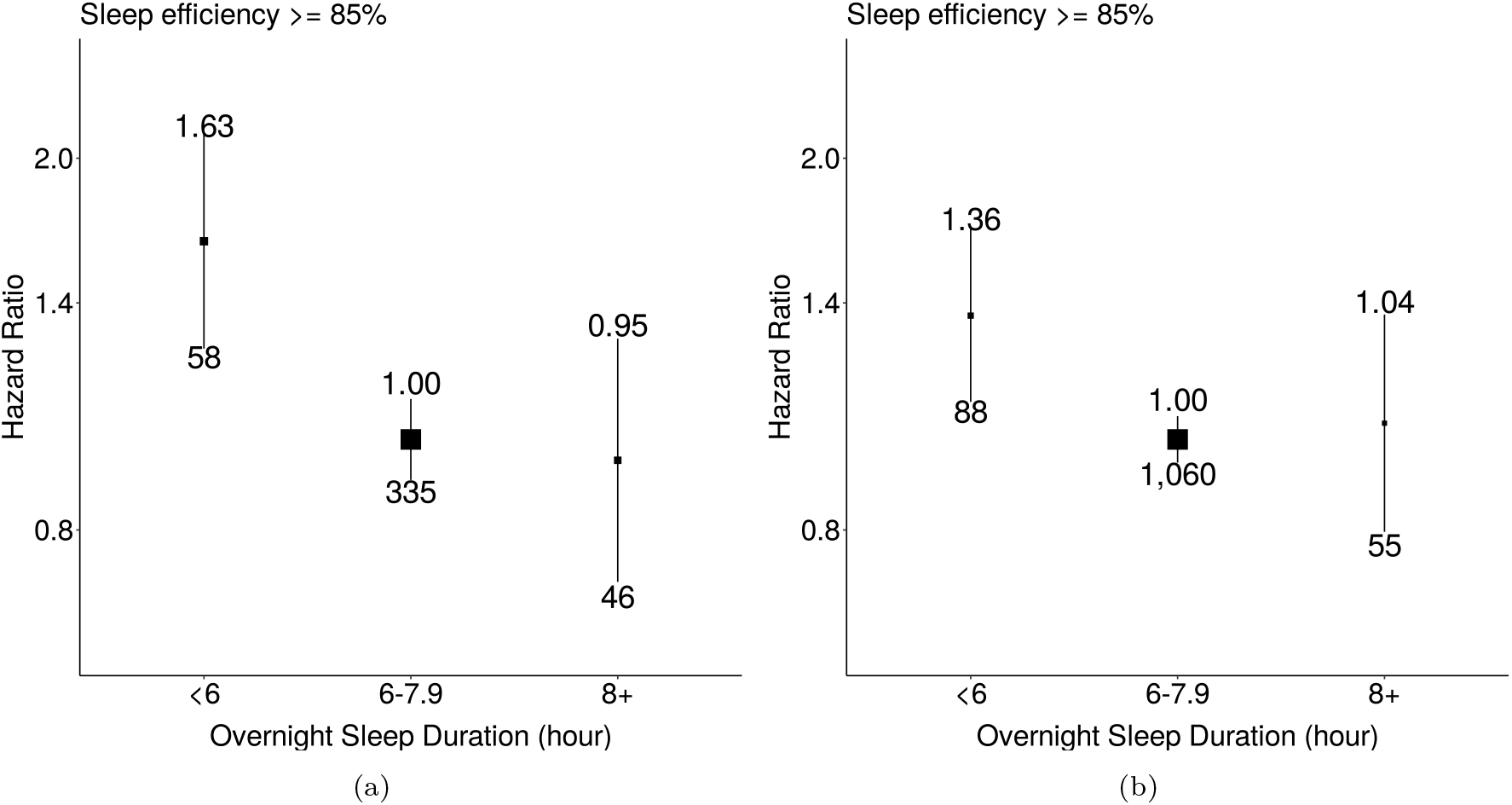
Associations of overnight sleep duration with all-cause mortality for groups with low and high sleep efficiency additionally adjusted for body mass index. The model used 1,642 events among 62,214 participants. We used age as the timescale and adjusted for sex, ethnicity, Townsend Deprivation Index of baseline address (split by quarter in the study population), educational qualifications, smoking status, alcohol consumption (Never, <3 times/week, 3+ times/week), overall activity (measured in milli-gravity units). Areas of squares represent the inverse of the variance of the log risk. The I bars denote the 95% confidence interval for the floated risks.

**Figure 25:**
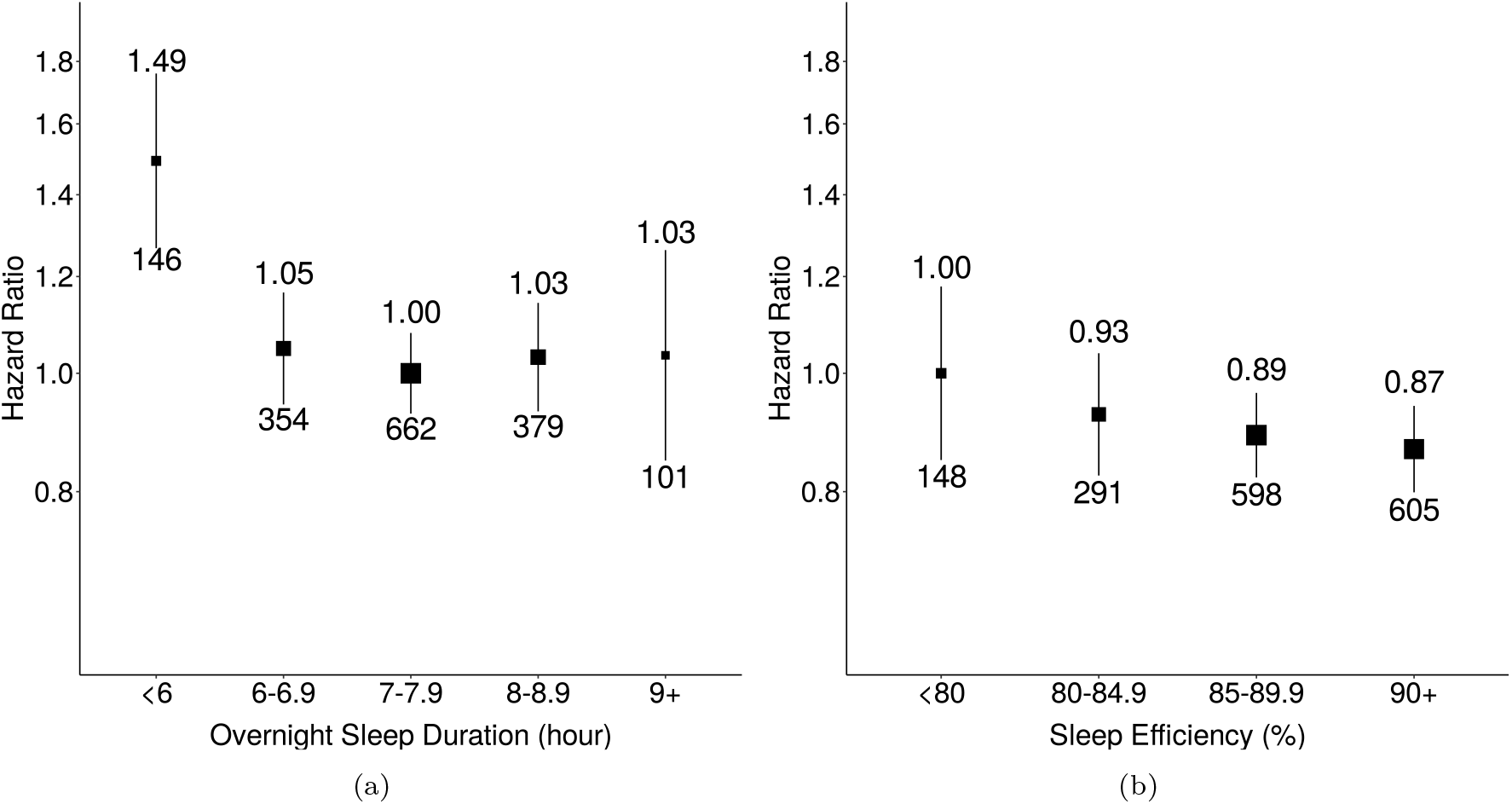
Associations of overnight sleep duration (a) and sleep efficiency (b) with all-cause mortality additionally adjusted for body mass index. The model used 1,642 events among 62,214 participants. We used age as the timescale and adjusted for sex, ethnicity, Townsend Deprivation Index of baseline address (split by quarter in the study population), educational qualifications, smoking status, alcohol consumption (Never, <3 times/week, 3+ times/week), overall activity (measured in milli-gravity units), and body mass index. Areas of squares represent the inverse of the variance of the log risk. The I bars denote the 95% confidence interval for the floated risks.

##### 8.3.2. Sensitivity analysis for overnight sleep duration

**Figure 26:**
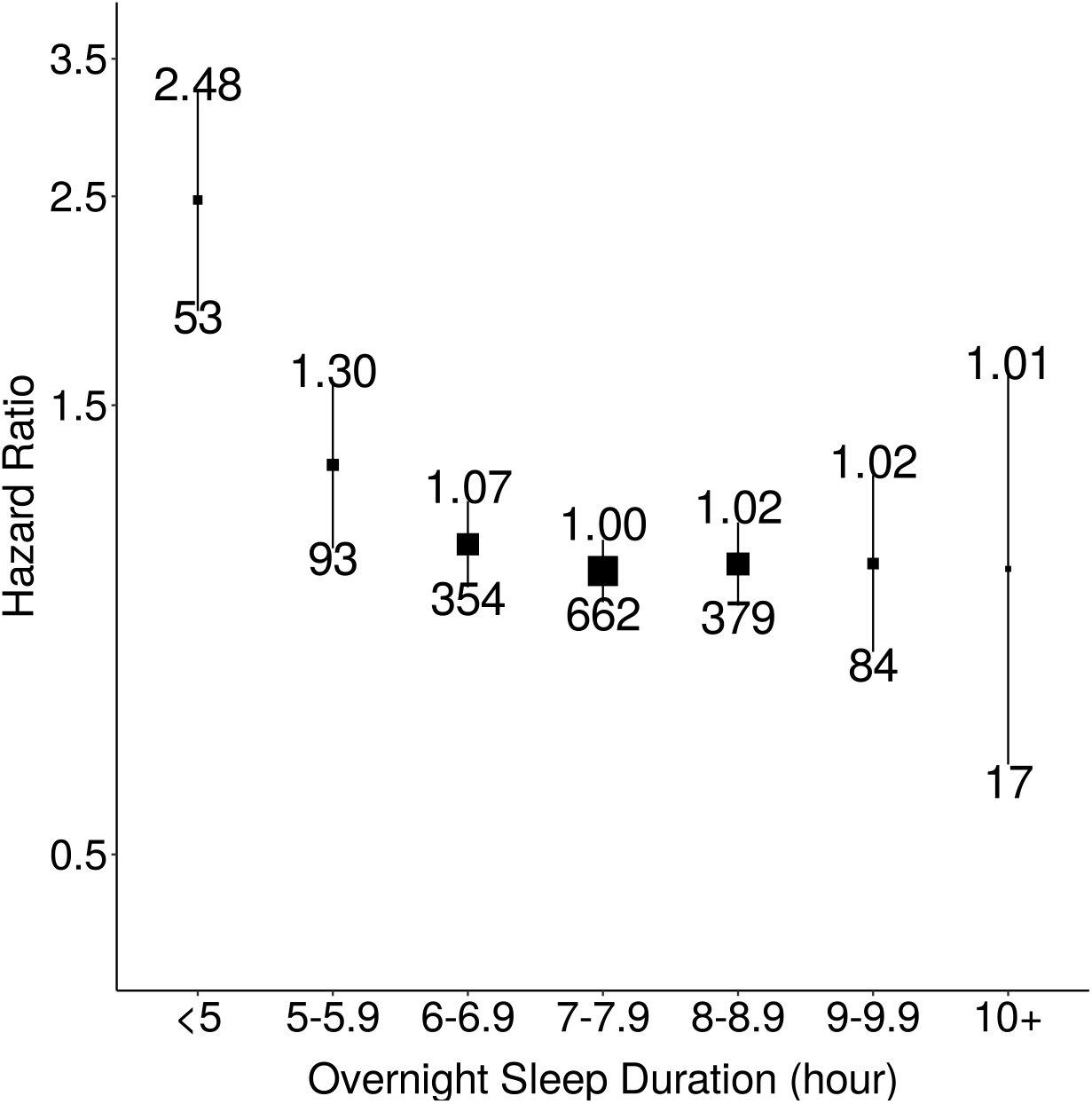
Associations of device-measured overnight sleep duration and all-cause mortality with greater granularity. The model used 1,642 events among 62,214 participants. We used age as the timescale and adjusted for sex, ethnicity, Townsend Deprivation Index of baseline address (split by quarter in the study population), educational qualifications, smoking status, alcohol consumption (Never, <3 times/week, 3+ times/week), and overall activity (measured in milli-gravity units). Areas of squares represent the inverse of the variance of the log risk. The I bars denote the 95% confidence interval for the floated risks.

